# Comparing natural language processing representations of disease sequences for prediction in the electronic healthcare record

**DOI:** 10.1101/2023.11.16.23298640

**Authors:** Thomas Beaney, Sneha Jha, Asem Alaa, Alexander Smith, Jonathan Clarke, Thomas Woodcock, Azeem Majeed, Paul Aylin, Mauricio Barahona

## Abstract

Natural language processing (NLP) is increasingly being applied to obtain unsupervised representations of electronic healthcare record (EHR) data, but their performance for the prediction of clinical endpoints remains unclear. Here we use primary care EHRs from 6,286,233 people with Multiple Long-Term Conditions in England to generate vector representations of sequences of disease development using two input strategies (212 disease categories versus 9,462 diagnostic codes) and different NLP algorithms (Latent Dirichlet Allocation, doc2vec and two transformer models designed for EHRs). We also develop a new transformer architecture, named EHR-BERT, which incorporates socio-demographic information. We then compare use of each of these representations to predict mortality, healthcare use and new disease diagnosis. We find that representations generated using disease categories perform similarly to those using diagnostic codes, suggesting models can equally manage smaller or larger vocabularies. Sequence-based algorithms perform consistently better than bag-of-words methods, with the highest performance for EHR-BERT.

## Introduction

Due to population aging and the effective management of many long-term conditions (LTCs), an increasing number of people are living with Multiple Long-Term Conditions (MLTC), a health state defined as the co-occurrence of two or more LTCs, and associated with a range of adverse health outcomes.^1,2^ Understanding the determinants and consequences of MLTC has become a priority for medical research and represents a significant shift from a focus on individual diseases to a focus on understanding the interactions of combinations of LTCs.^2–4^ Electronic healthcare records (EHRs), particularly those from primary care settings, which capture the history of a person’s diseases over time, provide a powerful resource for exploring these interactions. The growing use of EHRs in medical research has promoted the translation of methods which can handle such ‘big data’, such as those developed in natural language processing (NLP). Applied to the structured data in EHRs, the temporally-ordered sequence of medical codes or diseases in a patient’s record can be viewed as analogous to the sequence of words in a sentence or document.^5^ NLP approaches can be used to generate representations of patients based on their sequence of diseases;^6–8^ more recent transformer architectures can also be ‘fine-tuned’ to optimise the learned representation for prediction of clinical endpoint, such as next disease prediction.^5,9^

In healthcare settings, there are advantages to learning a single unsupervised representation of a patient based on their characteristics and diseases, rather than fine-tuning a different model for every outcome of interest. For example, segmentation methods aim to cluster patients for risk stratification, based on measures of similarity derived from demographic and clinical criteria; if one representation performs well across a range of outcomes, then only one representation and clustering could be trained and implemented, reducing complexity.^10^ However, there is little comparative research of the unsupervised representations of a person’s disease history generated by different NLP methods, and how these compare to standard epidemiological approaches in prediction of clinical outcomes. Furthermore, the optimal input strategies when translating methods designed for natural language to structured healthcare data remain uncertain. When analysing EHR data, a common approach is for clinical experts to manually group individual diagnosis codes (for example represented by ICD or SNOMED ontologies), into disease categories, to reduce the number of inputs into a model. This process is time-consuming, may be subjective, and risks discarding relevant information captured in a more specific code. If use of individual diagnosis codes alone can perform as well, or better than disease groupings, this might reduce the need for clinically derived categories as inputs for predictive models.

In this study, we compare different patient representations generated from widely used bag-of-words and sequence-based NLP methods: Latent Dirichlet Allocation (LDA), doc2vec and two transformer architectures designed for EHR data: Med-BERT and BEHRT.^5,9^ We also develop a new transformer architecture EHR-BERT, which includes additional demographic information. For each model, we compare as inputs using disease categories versus using a larger vocabulary of individual diagnostic codes. We then evaluate the performance of these unsupervised patient representations as inputs to a predictive model (logistic classifier) for clinically relevant outcomes over one year: mortality, emergency department (ED) attendance, emergency hospital admission, attendance with a condition or a new diagnosis of a condition.

## Results

### Data description

A total of 6,286,233 patients registered to GP practices in the Clinical Practice Research Datalink (CPRD) on 1^st^ January 2015 with two or more LTCs were eligible for inclusion (see Figure 1). Characteristics of the eligible population are displayed in Table 1. The mean (SD) age of the population was 53.8 (18.2) years, with slightly more females (53.1%) compared to males (46.9%). The majority were of White ethnicity (86.2%), followed by South Asian (5.9%) and Black (3.5%). There was a roughly even spread across the ten deciles of socioeconomic deprivation as measured by the Index of Multiple Deprivation (IMD), but with slightly more in the five most deprived deciles (52.3%).

**Figure 1:**
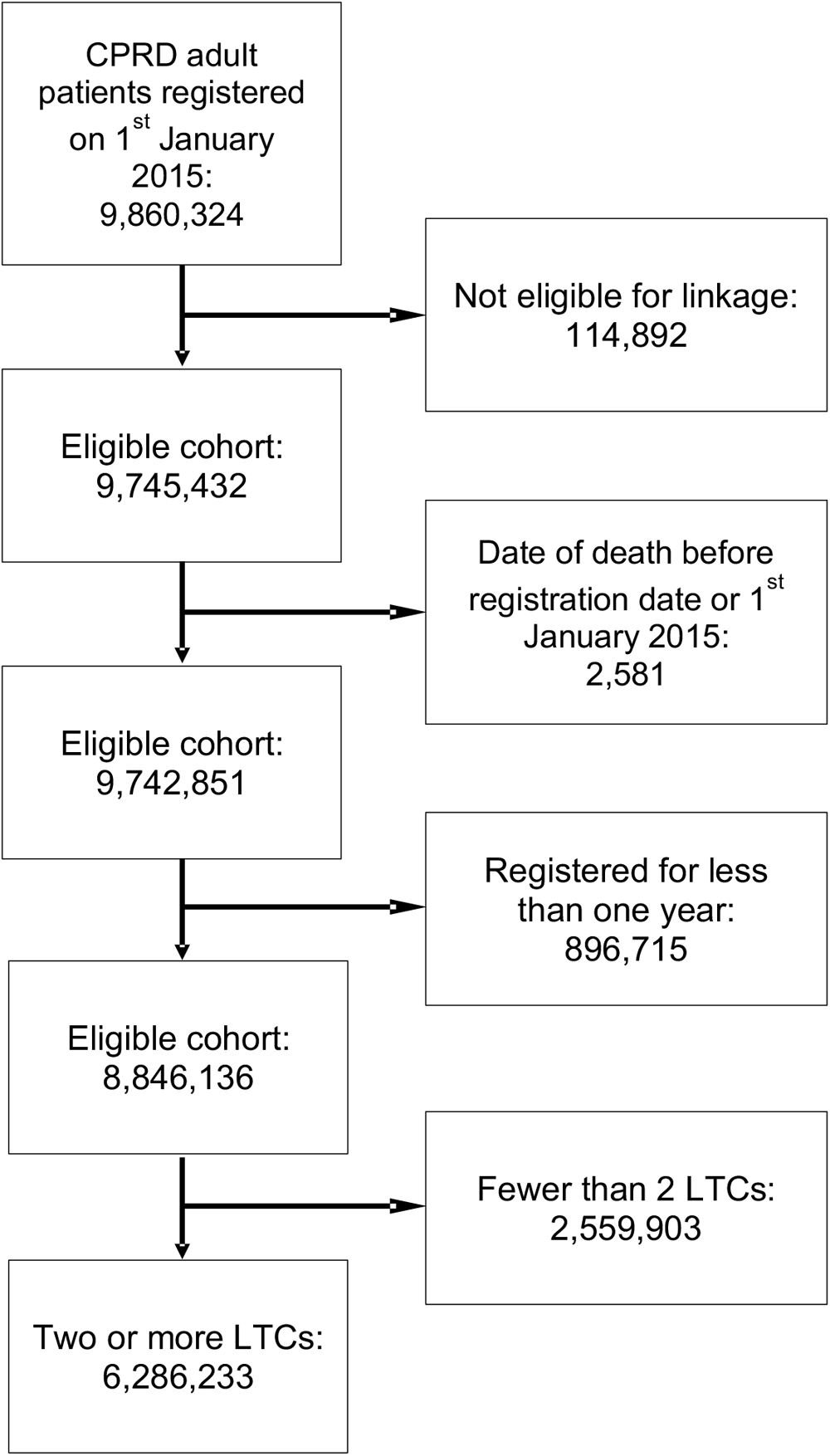
Flow chart of patients included in the study for generating embeddings.

**Table 1:**
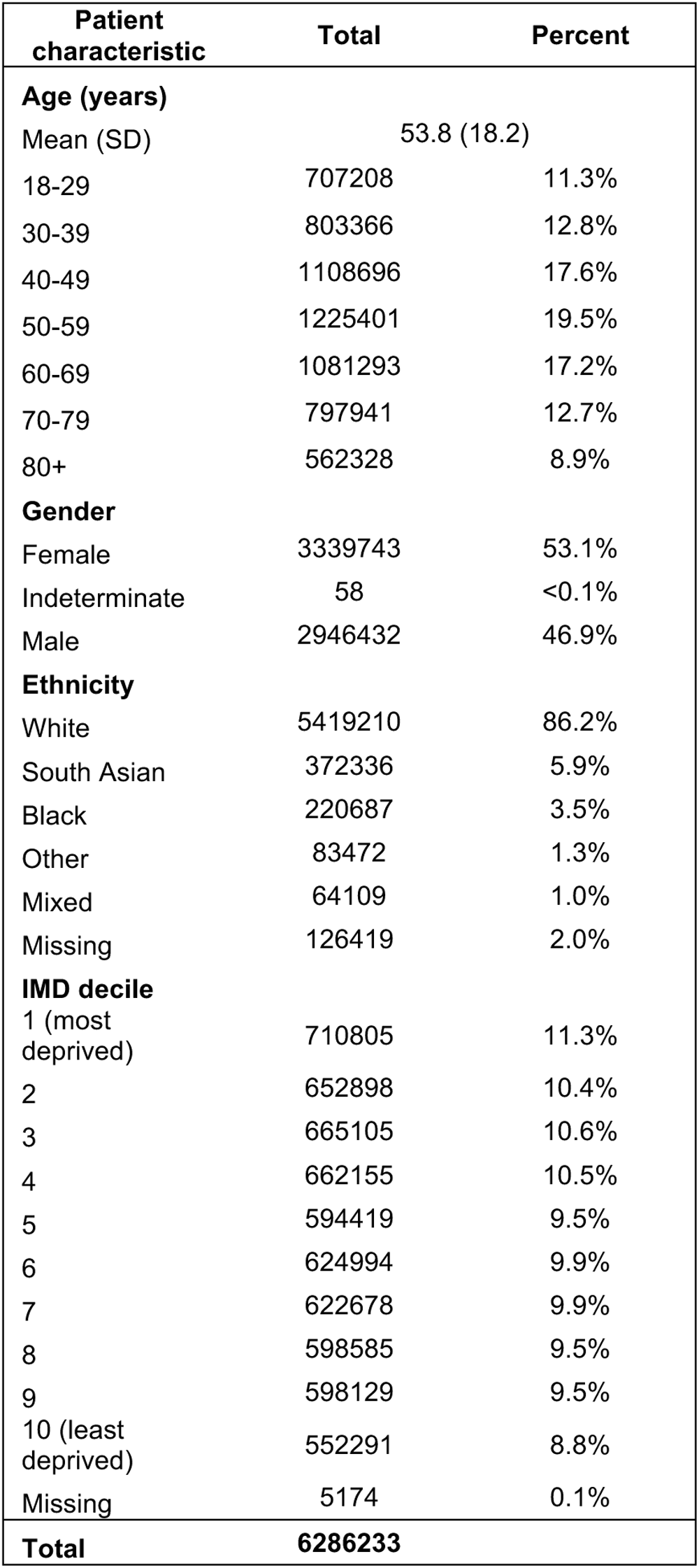
characteristics of the patient cohort (N = 6,286,233)

| Patient characteristic | Total | Percent |
| --- | --- | --- |
| **Age (years)** |  |  |
| Mean (SD) | 53.8 (18.2) |  |
| 18-29 | 707208 | 11.3% |
| 30-39 | 803366 | 12.8% |
| 40-49 | 1108696 | 17.6% |
| 50-59 | 1225401 | 19.5% |
| 60-69 | 1081293 | 17.2% |
| 70-79 | 797941 | 12.7% |
| 80+ | 562328 | 8.9% |
| **Gender** |  |  |
| Female | 3339743 | 53.1% |
| Indeterminate | 58 | <0.1% |
| Male | 2946432 | 46.9% |
| **Ethnicity** |  |  |
| White | 5419210 | 86.2% |
| South Asian | 372336 | 5.9% |
| Black | 220687 | 3.5% |
| Other | 83472 | 1.3% |
| Mixed | 64109 | 1.0% |
| Missing | 126419 | 2.0% |
| **IMD decile** |  |  |
| 1 (most deprived) | 710805 | 11.3% |
| 2 | 652898 | 10.4% |
| 3 | 665105 | 10.6% |
| 4 | 662155 | 10.5% |
| 5 | 594419 | 9.5% |
| 6 | 624994 | 9.9% |
| 7 | 622678 | 9.9% |
| 8 | 598585 | 9.5% |
| 9 | 598129 | 9.5% |
| 10 (least deprived) | 552291 | 8.8% |
| Missing | 5174 | 0.1% |
| **Total** | **6286233** |  |

### Generating patient representations

We generated vector representations of patients’ diseases using different NLP methods, comparing diseases versus Medcodes as inputs (Figure 2). Using LDA, we identified an optimal number of topics of 70 for diseases as inputs and 100 for Medcodes as inputs, based on the lowest perplexity score (see Supplementary Figures S2 and S3). With doc2vec, we found the Distributed Bag Of Words (DBOW) algorithm to perform better than the Distributed Memory (DM) algorithm in assigning patients with the same sequence as similar to each other (Supplementary Tables S3 and S4) and selected optimal models generating embeddings of size 100 for both disease and Medcode inputs. Finally, we created embeddings using the three transformer models, Med-BERT, BEHRT and EHR-BERT, each trained for 100 epochs.

**Figure 2:**
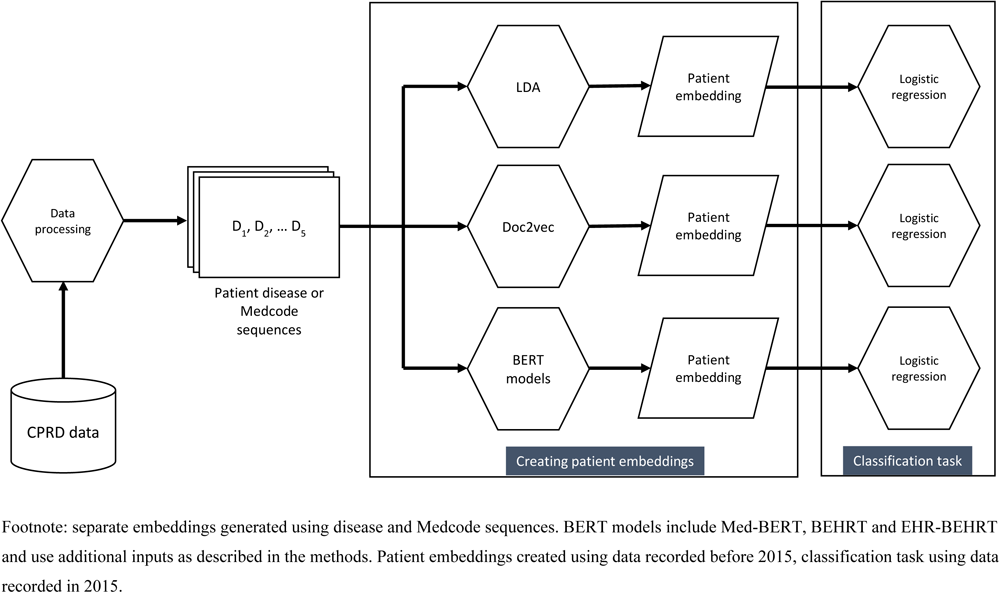
Analysis pipeline for generating patient embeddings and predicting outcomes.

### Evaluation of embedding performance

We evaluated the performance of the patient embeddings generated from each algorithm as inputs into a logistic classifier for predicting clinical outcomes over 12 months, including demographic covariates in the models. We compared these models to methods using binary indicators for having a disease (‘Binary disease indicators – unique’), using the count of codes for each disease (‘Disease frequency counts’), or using the demographic covariates alone as inputs.

We first assessed the performance, as measured by AUC and APS, of the patient embeddings generated from each algorithm in predicting mortality, ED attendance and emergency admissions (Figure 3). The embeddings tended to perform well at predicting mortality, but relatively poorly on predicting any ED attendance. Across all endpoints, embeddings generated by EHR-BERT performed best, followed by those from BEHRT, with use of binary disease indicators performing similarly to Med-BERT. The predictive model using the count of diagnosis codes for each disease (‘Disease frequency counts’) had the second lowest performance for all three endpoints, ahead only of models using sociodemographic covariates alone.

**Figure 3:**
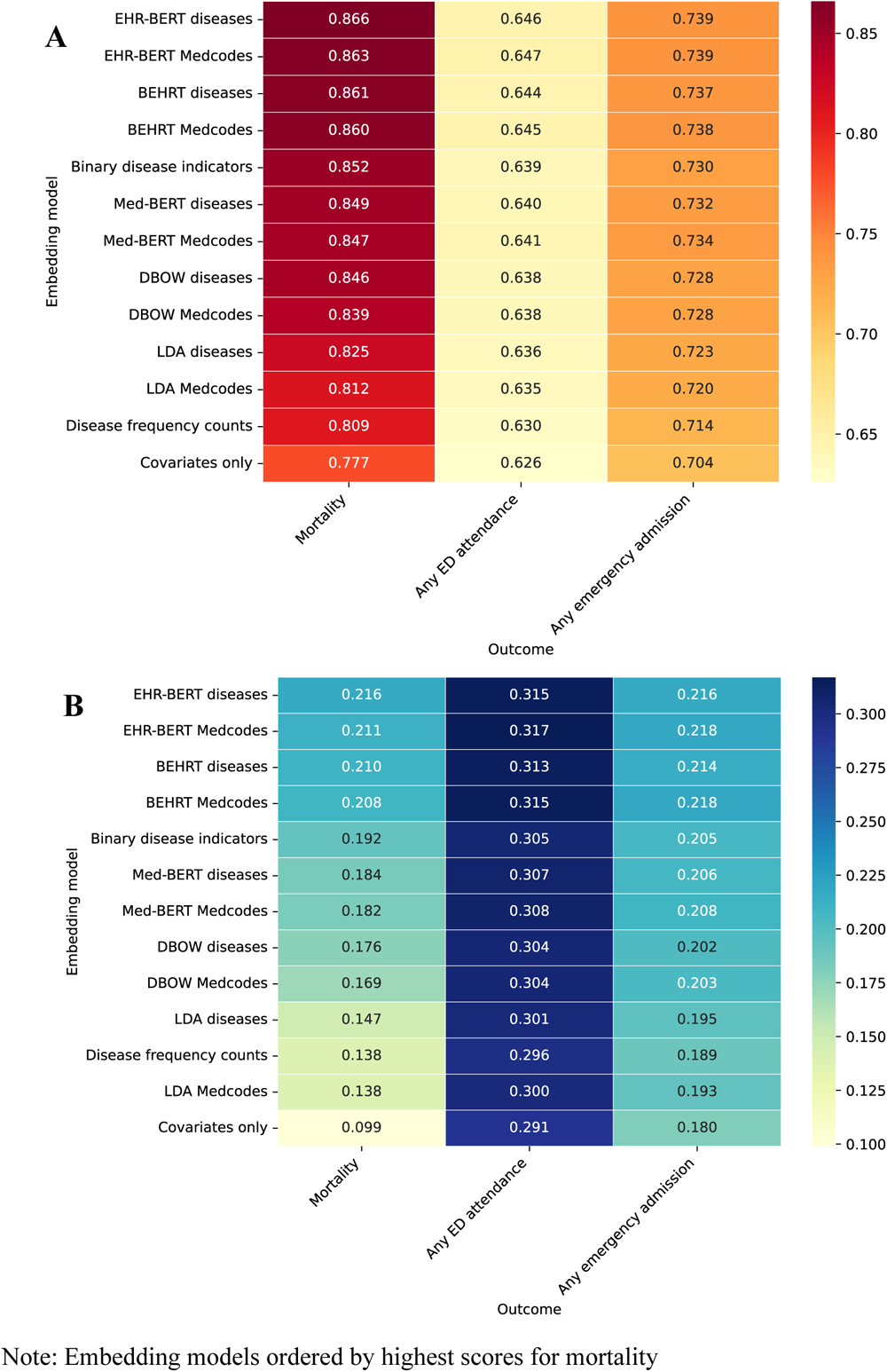
Model ROC-AUC (panel A) and APS (panel B) for different embedding models for prediction of mortality, emergency department attendances and emergency admissions within 12 months.

**Figure 4:**
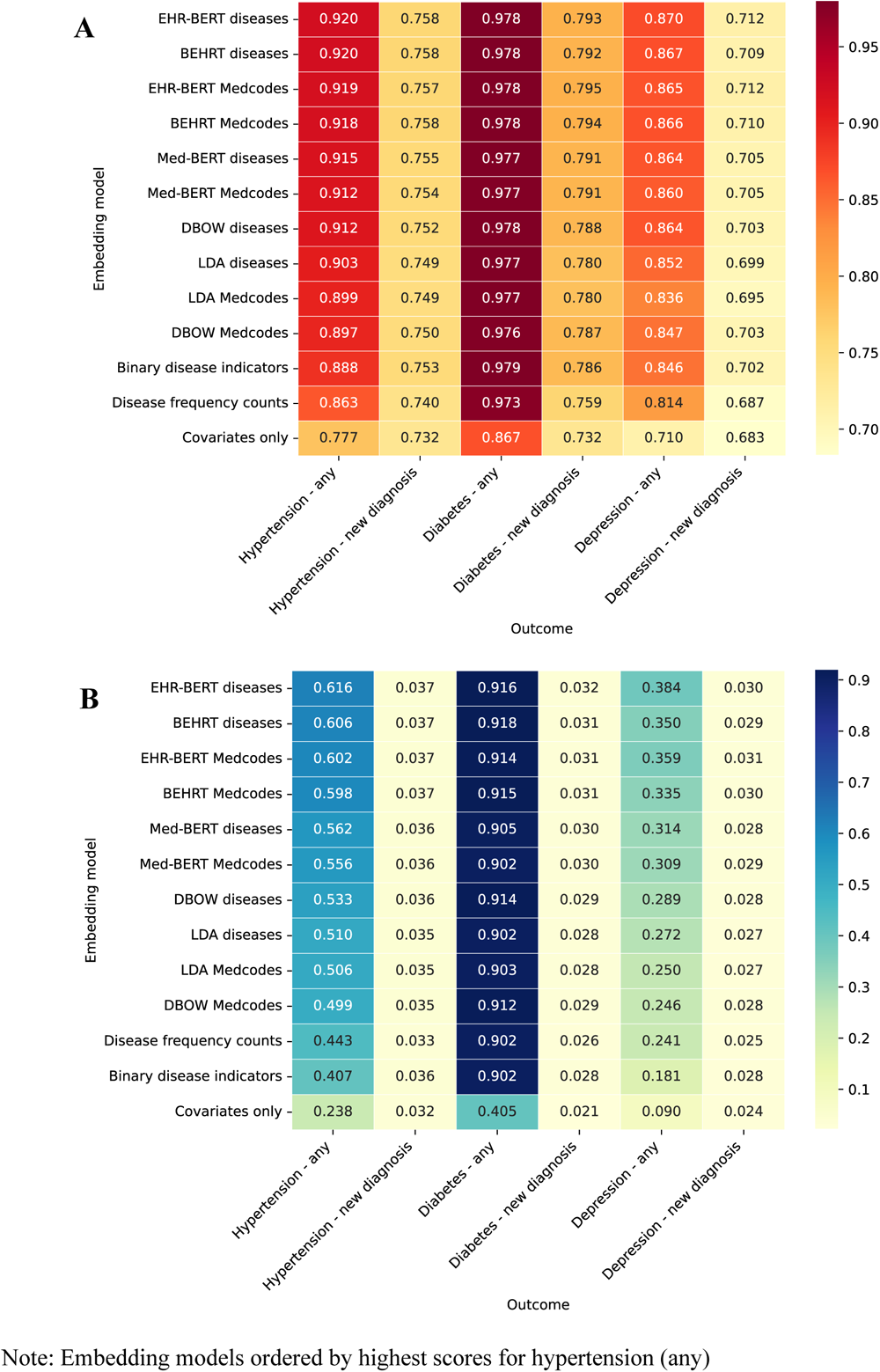
Model ROC-AUC (panel A) and APS (panel B) for different embedding models across for prediction of any versus new diseases developed within 12 months.

We next assessed the performance of the embeddings in predicting any attendance with hypertension, diabetes or depression over 12 months, versus models predicting new incidences of these diseases amongst patients not already diagnosed. For all diseases, the embeddings were better at predicting any occurrence than predicting newly incident diseases. Embeddings generated by EHR-BERT again performed best across all three diseases, followed by those from BEHRT and Med-BERT, and use of binary disease indicators performed comparatively worse. APS scores were consistently low for predicting new diseases.

A sensitivity analysis running the prediction models using the patient embeddings alone, without addition of the sociodemographic covariates showed worse overall performance, and similar relative differences with greater absolute differences between the embeddings (Supplementary Figures S5 and S6). However, for BEHRT, which includes age, and EHR-BERT, which includes sociodemographic variables in the MLM training, addition of sociodemographic covariates to the logistic regression classifier made little difference to the overall performance of the embeddings from these models.

## Discussion

Using a representative sample of six million people with MLTC, we compared different inputs and methods to learn representations of patients’ disease patterns as recorded in the primary care EHR and evaluated the performance of these representations in predicting a range of clinically relevant health outcomes. There are three key findings from our work. First, we directly demonstrated the predictive improvement of representations generated by models which incorporate information on disease sequence. Bag-of-words NLP methods performed comparatively poorly, and in general were worse than simply using binary disease indicators. BEHRT outperformed Med-BERT, highlighting the contextual importance of age in generating disease representations, while EHR-BERT improved on the performance of BEHRT, supplementing with knowledge of gender, ethnicity, socioeconomic deprivation, and calendar year. Second, we found that BEHRT and EHR-BERT performed well across all tasks, even without fine-tuning, indicating the ability of these unsupervised representations to capture latent information on sequence relevant to a range of health outcomes. Finally, across all embedding algorithms, we found little difference in the performance when using either the large vocabulary of Medcodes, or the smaller vocabulary of clinically categorised diseases.

### Implications for prediction models

Although transformer models performed best across all tasks, we found that using binary disease indicators generally performed well: better than both doc2vec and LDA in predicting mortality and healthcare attendance, and in predicting disease occurrence or incidence. This suggests that for many predictive tasks in clinical applications, particularly where model interpretability is important, there is little benefit of bag-of-words methods in prediction, particularly given the computational overhead. However, methods such as LDA, unlike doc2vec and transformer methods, may have an advantage in generating interpretable topic distributions which are more easily explainable.^11^ Notably, we found using the count of all repeated disease codes in the EHR performed particularly poorly and worse than methods using a single code occurrence, which suggests that crude code frequency in the EHR does not relate directly to disease severity and downstream risk. Previous work has highlighted the potential biases in code frequency in primary care data, which may relate to patient demographics, organisational policies, and coding incentives, rather than being an objective marker of a person’s health status.^12^ However, the better performance of BERT models, which also utilise recurrent codes, suggests that an ability of the attention mechanisms in these models to regulate the impact of repeated codes.

The similar performance of models using diseases (N=212) and Medcodes (N=9,462) suggests that the disease categories capture meaningful connections and use of the larger vocabulary does not add explanatory power. However, the manual grouping of codes into disease categories is a laborious process, that needs to be repeated as old codes are retired and new ones introduced. Our findings suggest that NLP models, as might be expected given their application to the large vocabulary found in natural language, can equally well manage with larger vocabulary, and extract relevant latent meaning. By extension, this indicates that inclusion of larger vocabularies of diagnosis codes, or use of other coded information, such as prescription data, may aid in prediction without the need for clinical categorisation.

Performance also varied according to the task. Prediction of any ED attendance was particularly poor, which likely relates to absence of other factors relating to attendance in our models, such as proximity and access to both primary care and ED departments and presentations with conditions not strongly related to MLTC, such as injuries.^13^ Although the embeddings generally performed well at predicting any attendance with a disease in the next 12 months, all performed poorly in predicting new diagnoses, with very low APS scores. In the original BEHRT paper (which also used CPRD data), the authors reported slightly higher AUC scores of 0.82, 0.81 and 0.88, compared with 0.76, 0.79 and 0.71 in our study, for hypertension, diabetes, and depression, respectively, which may relate to the additional benefit of fine-tuning beyond the MLM task that we employed.^9^ Solares *et al* (2021) also employed the disease embeddings used by BEHRT as weights to connect the inputs in a neural network, and found relatively higher precision scores (APS 0.15 for detecting hypertension).^8^ This might be explained by their use of a neural network architecture, which will allow for non-linear interactions between features, unlike logistic regression.

### Strengths and limitations

A strength of our study is the large and representative primary care dataset of six million people with MLTC in England.^14^ In contrast to studies using only secondary care data, primary care data contains the longitudinal history of a person’s health from birth to death, making them ideally suited to analyses of disease incidence. However, there are well-recognised biases in routinely collected data which impact on whether a code is recorded, dependent on it having taken place.^15^ First, there may be temporal delays in a code being entered, either due to late presentations by a patient, diagnostic uncertainty, or diagnostic delays, waiting until a firm diagnosis or referral is made, or correspondence is received from hospital.^16^ Second, the recurrence of codes in the medical record is dependent on factors including the GP practice, patient demographics, and disease-related factors, such as whether the disease is part of a quality assessment metric for practices.^12^ Previous research has highlighted that those in some ethnic groups, and living in areas of higher socioeconomic deprivation have a greater frequency of recurrent codes, which may result in models having more data, and therefore learning sequences better for some patients than others.^12^ In our study, we focussed on the performance across the population, but further work could explore the relative differences stratified by patient demographics, such as ethnicity.

A further strength of our study is the comparison of approaches applied to a single data source. However, a limitation of the large variety of methods trialled is the unavoidable difference in selection of the optimal hyperparameters between algorithms. With LDA, we used the perplexity score to determine the optimal number of topics, whereas for doc2vec, we used a metric based on creating a similar embedding for patients with identical disease sequences. This limitation is inherent to use of unsupervised methods, where there is no ground-truth for evaluating a learned embedding and represented a pragmatic approach. Nevertheless, it is likely that further hyper-parameter optimisation could improve upon the learned representations from LDA and doc2vec algorithms. Similarly, and in common with other machine learning approaches, BERT architectures include many hyperparameters which may affect performance. In our work, we used settings identified by other authors, rather than seek to evaluate the optimal configurations for our case, but this represents an avenue for further work. Furthermore, we used patient representations generated from the second-to-last hidden layer; further exploration of the optimal layer or combination of layers may lead to improved performance. However, previous literature applied to language has suggested only small differences between layer choice.^17,18^

Given our focus on the direct comparison of the embeddings as inputs, we used only logistic regression, given its popularity in epidemiological research, interpretability, and relatively few hyperparameters for tuning. Use of different classification algorithms, such as kernel-based algorithms, multi-layer perceptron or gradient boosting algorithms may result in better predictive performance of the embeddings, particularly given the likelihood of non-linear interactions between features.^6^ However, these algorithms also require a significant amount of optimisation, with hyperparameters that are likely to vary both by the embedding input and the outcome, and would complicate comparative interpretation.

## Conclusion

Comparing different NLP algorithms for representing patients’ disease patterns, we found that approaches using sequences performed significantly better in predicting clinical endpoints than methods using co-occurrence alone. Unsupervised patient representations from transformer architectures performed well across tasks without the need for fine-tuning, indicating the potential for learning multi-purpose representations which could be used in future for segmentation and risk stratification.

## Methods

### Data sources

We used CPRD Aurum, a nationally representative EHR dataset which includes routinely collected healthcare data from General Practices in England.^14^ Our data extract includes all patients aged 18 years or over, marked as ‘research acceptable’ (a data quality marker defined by CPRD, such as including a valid date of birth and registration date^19^) and registered to a GP practice on 1^st^ January 2015. We included all patients with two or more LTCs (defined below) eligible for linkage to secondary care data and those registered to a practice for at least one year to ensure sufficient time for data input.^20^ Cleaning rules for demographic data including age, gender and ethnicity are given in the Supplementary Information p.2-3. As a marker of socioeconomic deprivation, data were linked to deciles of the 2019 IMD of a patient’s geographical area of residence.^21^ Secondary care data was sourced from Hospital Episode Statistics (HES) data and data on death registrations from the Office for National Statistics, both linked to the Aurum dataset by CPRD.^22^ As CPRD Aurum data also includes a marker of mortality, any differences in the dates between sources were reconciled these using a modification of the algorithm recommended by Delmestri and Prieto-Alhambra (2020) (see Supplementary Information p.3).^23^

### Disease definition and sequence construction

Coded data entered during clinical encounters are stored in CPRD as numeric ‘Medcodes’. We included 9,462 Medcodes representing a group of 212 diseases defined in our previous study as LTCs.^24–26^ For each patient, we constructed a sequence of diagnostic codes ordered by date of occurrence representing the full history of these Medcodes from birth until 1^st^ January 2015. All retrospective clinical data for each patient is included in CPRD (i.e., prior to the study start date), where clinical diagnoses may be back-dated to the date of diagnosis rather than the date of data-entry. We created two separate sequences: the first, ‘Disease’, sequence using the disease categories (vocabulary size = 212), and the second, ‘Medcodes’ sequence using the individual Medcodes (vocabulary size= 9,462). Given our inclusion of patients with two or more LTCs, all sequences contained at least two diseases or Medcodes.

### Patient representation methods

We applied a range of methods for deriving document representations (embeddings) in NLP. For each method, we ran separate models using the disease sequences or Medcode sequences (see Figure 2 for our analysis pipeline).

### Latent Dirichlet Allocation

LDA is a generative probabilistic model which applies a distribution of topics over each document, assuming topics are drawn from a Dirichlet distribution.^27^ Details of our model set-up and choice of priors are given in the Supplementary Information. To select the optimal number of topics, we divided our data into an 80:20 train-test split and selected the number of topics resulting in the lowest value of the perplexity score on a test set not seen during training (see Supplementary Information, p.4-6).^28^ Using the optimal number of topics, we then repeated the LDA algorithm on the full dataset to generate topic distributions for the full patient cohort.

### Doc2vec

Doc2vec is an extension of the word2vec algorithms, which directly learns vector representations of text sequences ranging in length from sentences to documents.^29^ Further details of our hyperparameter tuning are given in the Supplementary Information, based on our previous work using word2vec.^26^ We found that the default learning rate and epochs for text produced poor representations of the patient embeddings, when using diseases, rather than Medcodes, as inputs. To evaluate the performance of doc2vec models using different learning rates and epochs, we made use of patients with identical sequences in the record, who we would expect to have a similar embedding vector (further details given in the Supplementary Information, p.7-11). We compared both DBOW and DM algorithms for both disease and Medcode sequences. The DM model learns features by approximating individual words (diseases or Medcodes in our case) using the surrounding context and the DBOW model samples a random word from a patient sequence to approximate the surrounding context. The former approach takes order into consideration, but is more likely to learn similar patterns with smaller lengths of sequence.^29^

### Transformer models

We compared Med-BERT and BEHRT, two recent transformer-based architectures designed for coded EHR data. Med-BERT used as input 82,000 ICD-9 and ICD-10 codes from hospital EHR data for 20 million patients,^5^ whereas BEHRT grouped codes from 1.6 million patients’ primary care EHR data into 301 disease categories, and supplemented training with information on patient age at each code occurrence.^9^ Although the original BERT implementation included an additional pre-training step with a next-sentence prediction task, further studies have suggested improvement in downstream language tasks using masked-language-modelling (MLM) alone.^30^ We therefore used MLM alone (as done in BEHRT) and did not conduct the further pre-training step performed in Med-BERT. The MLM approach in a Transformer architecture enables the model to learn a deep, bidirectional representation of an entire sequence. The attention mechanism, which is a mainstay of the Transformer-based architectures, allows the model to capture longer range of dependencies between the EHR codes, leading to better capturing of context. To better enable direct comparison between models, we made some changes to the default implementations and fixed the minimum sequence length per patient to two, and the maximum sequence length to 128, which accounted for the full sequence for 97.3% of patients (Table 2). Sequences longer than 128 were truncated, retaining the most recent 128 codes.

**Table 2:**
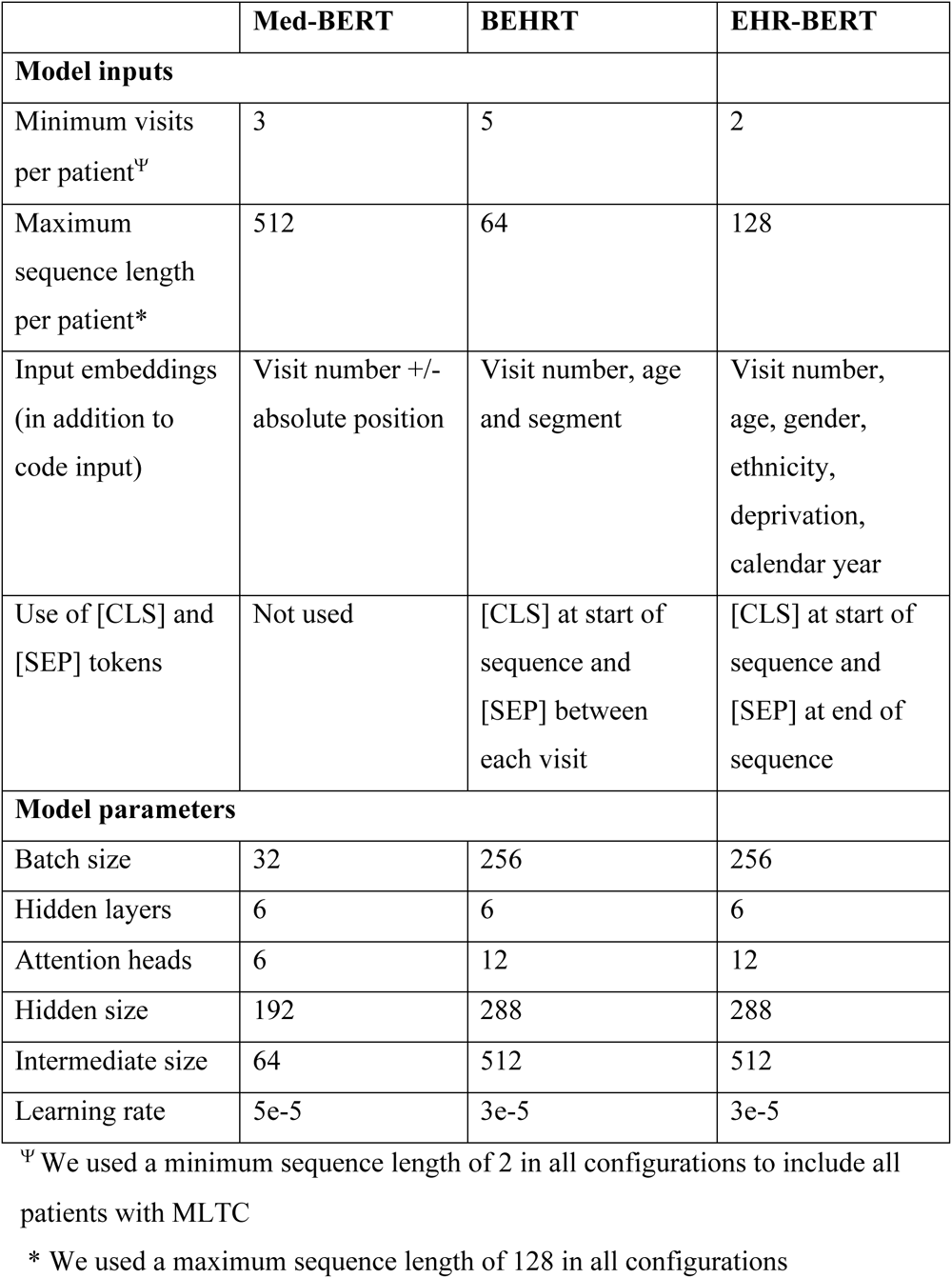
Model inputs and parameters for Med-BERT, BEHRT and EHR-BERT architectures.

We also developed a new transformer architecture, which we call EHR-BERT. EHR-BERT adapts and extends the BEHRT model to include additional sociodemographic factors known to be associated with code frequency in the EHR.^12^ We added embedding layers for gender, ethnicity, socioeconomic deprivation decile and calendar year of the observation (Table 2 and Supplementary Information Figure S4). Compared to BEHRT, we removed the segment token, and the [SEP] tokens between visit, as most visits contained only one code, and we believed that visit number should suffice to capture sequential visit information.

The six models (three BERT variants, each with two separate model inputs (diseases or Medcodes) were trained on the full study cohort for 100 epochs, as used in BEHRT. To generate patient embeddings, we averaged the second-to-last hidden layer of each of the contextualised disease embeddings in a patient’s sequence, resulting in a vector equal in length to the hidden size.^17,31^

### Evaluation

We evaluated the performance of the embeddings in predicting the following binary outcomes measured in the 12 months after 1^st^ January 2015 (the final date used for generating the embeddings), using data not seen during training:

1. Mortality (of patients aged ≥60 years on 1^st^ January 2015)
2. Any emergency department (ED) attendance
3. Any emergency hospital admission
4. Any attendance with a coded diagnosis code for:

a. Hypertension
b. Diabetes (Type 1 or Type 2, or unspecified)
c. Depression
5. A new diagnosis of:

a. Hypertension
b. Diabetes (Type 1 or Type 2, or unspecified)
c. Depression

The three diseases were selected based on relatively high frequency in the dataset (Supplementary Information Table S2) and as important for early detection. For all outcomes, we excluded 305,142 patients who deregistered during the 12-month period, to ensure equal follow-up time (total population = 5,981,091). For all outcomes except mortality, we excluded the 85,260 patients who died during the 12-month follow-up (total population = 5,895,831). For prediction of new diagnoses only, we additionally excluded any patient from analysis who already had the disease of interest diagnosed by 1^st^ January 2015.

As a comparison to a standard epidemiological approach without information on co-occurrence or sequence, we constructed a ‘Binary disease indicator’ embedding where each of the 212 disease categories was represented as a binary feature (present or not present). We also constructed a ‘Disease frequency count’ embedding where each feature represented the number of times each disease appeared in the record for the patient.

To compare performance of different representations, we used them as inputs (features) into a logistic regression classifier, running separate models for each of the outcomes described above. Logistic regression models were trained with four-fold cross-validation, with L2 penalisation (regularization strength of 1) and maximum iterations of 100. We experimented with smaller and larger values of the regularization parameter (0.01, 0.1 and 10) but found 1 to be optimal. We included age, gender, ethnicity and deprivation decile, categorised as in Table 1. As a sensitivity analysis, we compared model performance using the patient representations alone, without inclusion of covariates.

Models were evaluated on the test sets using the Receiver Operating Characteristic Area Under the Curve (ROC-AUC) score and Average Precision Score (APS), which is the area under the precision-recall curve and gives a better indication of model performance for predicting the positive class.^32^ As a sensitivity analysis for the Med-BERT, BEHRT and EHR-BERT models, we compared the performance at 10, 50 and 100 epochs, and selected 100 epochs as the best performing (see Supplementary Information Tables S5 and S6).

### Implementation

We used Python version 3.10.9 and Pandas version 2.0.3 for data manipulation and management.^33,34^ Skip-gram, LDA and doc2vec were applied using the gensim library.^35^ The BEHRT model was implemented using the authors’ source code available on GitHub (https://github.com/deepmedicine/BEHRT). We implemented Med-BERT and EHR-BERT using the HuggingFace Transformer library;^36^ Med-BERT used the same architecture and hyperparameter settings described by the authors.^5^ Transformer models were run on an NVIDIA Quadro RTX 8000 GPU with 48GB RAM; each model took approximately ten days to run for 100 epochs. Codes, including the Medcode to disease mapping are available from https://tbeaney.github.io/MMclustering/

## Data Availability

This study uses patient data which is not publicly available but can be requested for users meeting certain requirements: https://cprd.com/research-applications. Codes, including the Medcode to disease mapping are available from https://tbeaney.github.io/MMclustering/

## Ethics

Data access to CPRD and ethical approval was granted by CPRD’s Research Data Governance Process on 28^th^ April 2022 (Protocol reference: 22_001818) and with linkage to HES data on 6^th^ March 2023 (Protocol reference: 22_002481).

## Acknowledgements

This research is funded through a clinical PhD fellowship awarded to TB from the Wellcome Trust 4i programme at Imperial College London. Data management was provided by the Big Data and Analytical Unit (BDAU) at the Institute of Global Health Innovation (IGHI), Imperial College London. We are grateful for the support of the NIHR Imperial Biomedical Research Centre. JC acknowledges support from the Wellcome Trust (215938/Z/19/Z). TW, AM and PA acknowledge support from the National Institute for Health and Care Research (NIHR) Applied Research Collaboration Northwest London. MB acknowledges support from EPSRC grant EP/N014529/1 supporting the EPSRC Centre for Mathematics of Precision Healthcare. The views expressed in this publication are those of the authors and not necessarily those of the NHS, the NIHR, the Wellcome Trust or the Department of Health and Social Care.

## References

1. Johnston, M. C., Crilly, M., Black, C., Prescott, G. J. & Mercer, S. W. Defining and measuring multimorbidity: a systematic review of systematic reviews. European Journal of Public Health 29, 182–189 (2019).

2. Pearson-Stuttard, J., Ezzati, M. & Gregg, E. W. Multimorbidity—a defining challenge for health systems. The Lancet Public Health 4, e599–e600 (2019).

3. Whitty, C. J. M. & Watt, F. M. Map clusters of diseases to tackle multimorbidity. Nature 579, 494–496 (2020).

4. The Academy of Medical Sciences. Multimorbidity: a priority for global health research. Academy of Medical Sciences (2018).

5. Rasmy, L., Xiang, Y., Xie, Z., Tao, C. & Zhi, D. Med-BERT: pretrained contextualized embeddings on large-scale structured electronic health records for disease prediction. *npj Digit*. Med. 4, 1–13 (2021).

6. Choi, E., Schuetz, A., Stewart, W. F. & Sun, J. Medical Concept Representation Learning from Electronic Health Records and its Application on Heart Failure Prediction. Preprint at http://arxiv.org/abs/1602.03686 (2017).

7. Choi, E. et al. Multi-layer representation learning for medical concepts. Proceedings of the ACM SIGKDD International Conference on Knowledge Discovery and Data Mining 13-17-Augu, 1495–1504 (2016).

8. Solares, J. R. A. et al. Transfer Learning in Electronic Health Records through Clinical Concept Embedding. 1–14 (2021).

9. Li, Y. et al. BEHRT: Transformer for Electronic Health Records. Sci Rep 10, 7155 (2020).

10. Yan, S., Kwan, Y. H., Tan, C. S., Thumboo, J. & Low, L. L. A systematic review of the clinical application of data-driven population segmentation analysis. BMC Medical Research Methodology 18, 121 (2018).

11. Lannou, E. L. et al. Clustering of patient comorbidities within electronic medical records enables high-precision COVID-19 mortality prediction. medRxiv 2021.03.29.21254579 (2021).

12. Beaney, T. et al. Identifying potential biases in code sequences in primary care electronic healthcare records: a retrospective cohort study of the determinants of code frequency. BMJ Open 13, e072884 (2023).

13. Giebel, C. et al. What are the social predictors of accident and emergency attendance in disadvantaged neighbourhoods? Results from a cross-sectional household health survey in the north west of England. BMJ Open 9, e022820 (2019).

14. Wolf, A. et al. Data resource profile: Clinical Practice Research Datalink (CPRD) Aurum. International Journal of Epidemiology 48, 1740–1740g (2019).

15. Verheij, R. A., Curcin, V., Delaney, B. C. & McGilchrist, M. M. Possible Sources of Bias in Primary Care Electronic Health Record Data Use and Reuse. J Med Internet Res 20, e185 (2018).

16. Ford, E. et al. What evidence is there for a delay in diagnostic coding of RA in UK general practice records? An observational study of free text. BMJ Open 6, e010393 (2016).

17. Devlin, J., Chang, M. W., Lee, K. & Toutanova, K. BERT: Pre-training of deep bidirectional transformers for language understanding. NAACL HLT 2019-2019 Conference of the North American Chapter of the Association for Computational Linguistics: Human Language Technologies - Proceedings of the Conference 1, 4171–4186 (2019).

18. Frequently Asked Questions — bert-as-service 1.6.1 documentation. https://bert-as-service.readthedocs.io/en/latest/section/faq.html#how-large-is-a-sentence-vector.

19. Herrett, E. et al. Data Resource Profile: Clinical Practice Research Datalink (CPRD). International Journal of Epidemiology 44, 827–836 (2015).

20. Lewis, J. D., Bilker, W. B., Weinstein, R. B. & Strom, B. L. The relationship between time since registration and measured incidence rates in the General Practice Research Database. Pharmacoepidemiology and Drug Safety 14, 443–451 (2005).

21. Ministry of Housing & Communities & Local Government. English indices of deprivation 2019. https://www.gov.uk/government/statistics/english-indices-of-deprivation-2019.

22. NHS Digital. Hospital Episode Statistics (HES). https://digital.nhs.uk/data-and-information/data-tools-and-services/data-services/hospital-episode-statistics.

23. Delmestri, A. & Prieto-Alhambra, D. CPRD GOLD and linked ONS mortality records: Reconciling guidelines. Int J Med Inform 136, 104038 (2020).

24. Kuan, V. et al. A chronological map of 308 physical and mental health conditions from 4 million individuals in the English National Health Service. The Lancet Digital Health 1, e63–e77 (2019).

25. Head, A. et al. Inequalities in incident and prevalent multimorbidity in England, 2004&#x2013;19: a population-based, descriptive study. The Lancet Healthy Longevity 2, e489–e497 (2021).

26. Beaney, T. et al. Identifying multi-resolution clusters of diseases in ten million patients with multimorbidity in primary care in England. 2023.06.30.23292080 Preprint at 10.1101/2023.06.30.23292080 (2023).

27. Blei, D. M., Ng, A. Y. & Jordan, M. I. Latent Dirichlet Allocation. Journal of Machine Learning Research 3, 993–1022 (2003).

28. Röder, M., Both, A. & Hinneburg, A. Exploring the Space of Topic Coherence Measures. in Proceedings of the Eighth ACM International Conference on Web Search and Data Mining 399–408 (Association for Computing Machinery, 2015). doi:10.1145/2684822.2685324.

29. Le, Q. & Mikolov, T. Distributed Representations of Sentences and Documents. in Proceedings of the 31st International Conference on Machine Learning 1188–1196 (PMLR, 2014).

30. Liu, Y. et al. RoBERTa: A Robustly Optimized BERT Pretraining Approach. Preprint at http://arxiv.org/abs/1907.11692 (2019).

31. Xiao, H. bert-as-service. (2018).

32. Davis, J. & Goadrich, M. The relationship between Precision-Recall and ROC curves. in Proceedings of the 23rd international conference on Machine learning 233–240 (Association for Computing Machinery, 2006). doi:10.1145/1143844.1143874.

33. The Python Language Reference. Python documentation https://docs.python.org/3/reference/index.html.

34. McKinney, W. Data Structures for Statistical Computing in Python. in 56–61 (2010). doi:10.25080/Majora-92bf1922-00a.

35. Řehůřek, R. & Sojka, P. Software Framework for Topic Modelling with Large Corpora. in Proceedings of the LREC 2010 Workshop on New Challenges for NLP Frameworks 45–50 (ELRA, 2010).

36. Wolf, T., et al. HuggingFace’s Transformers: State-of-the-art Natural Language Processing. Preprint at 10.48550/arXiv.1910.03771 (2020).

