## Supplementary Information for "Comparing natural language processing representations of disease sequences for prediction in the electronic healthcare record"

**Ethnicity data cleaning**

NHS healthcare data in England uses the same ethnic categories as the 2001 census, which categorise ethnicity into a 5-level and a 16-level grouping.^1^ We used the 5-level grouping using the ethnicity codes developed by Davidson *et al* (2021).^2^ We removed four codes from these list representing examination findings, as suggesting clinician observed findings rather than reported by the patient. We followed the algorithm from Mathur *et al* (2020) to identify ethnicity:^3^

1. Remove duplicate ethnicity codes recorded on the same date
2. Assign each code to the 5 and 16 level ethnicity categories
3. If only one ethnicity code, use single code to define ethnicity
4. If more than one code, use most frequently occurring ethnicity code
5. If more than one code with the same count, use the latest ethnicity code instead.

**Reconciliation of death recording**

Deaths are recorded in both CPRD and in the Office for National Statistics (ONS) mortality statistics, of which the latter are generally regarded as the gold standard for recording deaths, given the legal requirement for its reporting in England and Wales*.*^4^ However, the ONS figures represent only deaths recorded in England and Wales; the 7,524 deaths present only in CPRD may therefore reflect deaths that occurred outside of England and Wales, which would not be recorded in ONS but still represent genuine data. Delmestri and Prieto-Alhambra (2020) developed guidelines for reconciling the two sources of deaths.^5^ Although their study used CPRD GOLD, the process of death registration in GP practices contributing to CPRD Aurum is likely to be similar, making these recommendations applicable. We adapted their recommendations to reconcile the two sources of death, as follows:

1. Use CPRD date of death if this is the only source present.
2. Use ONS date of death if this is the only source present and the match rank equals 1 or 2.
3. If data are recorded in both sources, use ONS only if match rank equals 1 or 2, or CPRD if match rank 3 or more.

Here, the match rank refers to CPRD’s linkage criterion, with 1 and 2 corresponding to a match including the full date of birth to increase the robustness of the linkage.

**Table S1: Sequence lengths for each patient**

| **Sequence length** | **Number (%)** | |
| --- | --- | --- |
| Mean | 27.1 | |
| Median | 13 | |
| >32 | 1459380 | (23.22%) |
| >64 | 599344 | (9.53%) |
| >128 | 171646 | (2.73%) |
| >256 | 44415 | (0.71%) |
| Total | 6286233 | |

**Table S2: count of number of incident (newly diagnosed) diseases during one year of follow-up, for diseases with at least 5,000 occurrences**

| **Disease** | **Count** |
| --- | --- |
| Raised Total Cholesterol | 24321 |
| Raised LDL-C | 20267 |
| Raised Triglycerides | 18769 |
| Enthesopathy and synovial disorder | 16549 |
| Low HDL-C | 16523 |
| Obesity | 13000 |
| **Hypertension** | 12709 |
| Osteoarthritis (excl spine) | 12139 |
| Anxiety disorders | 11989 |
| **Depression** | 10587 |
| Dermatitis | 10030 |
| Gastro-oesophageal reflux disease | 9178 |
| Hearing loss | 8857 |
| **Diabetes Mellitus other or not specified*** | 8324 |
| Cataract | 7767 |
| **Type 2 Diabetes Mellitus*** | 7177 |
| Chronic Kidney Disease | 6063 |
| Allergic and chronic rhinitis | 5587 |
| Atrial Fibrillation | 5483 |
| Gastritis and duodenitis | 5461 |
| Diverticular Disease | 5396 |
| Alcohol Misuse | 5032 |

* Note diabetes diseases are grouped in analysis

**LDA**

We assumed asymmetric Dirichlet priors for both disease-topic and patient-topic distributions. Previous studies have suggested using asymmetric document-topic distributions produce more coherent topics, but found little difference in the choice of word-topic prior.^6,7^ In our case, our *a priori* expectation is that the disease-topic prior should be asymmetric, as we would not expect many diseases to occur in multiple topics (indeed, this would be an undesirable property). We used the *gensim* implementation of LDA which implements the online variational Bayes algorithm developed by Hoffman *et al* (2010), and estimates the values of the asymmetric priors.^8,9^

We estimated the number of passes and iterations required through the whole dataset using a sub-sample of the data. Given the computational load of calculating the evaluation metrics, we selected sub-samples of 0.1% and 1% of the data, and calculated convergence and perplexity metrics across a range of iterations and passes with a fixed number of 20 topics. Results from the 1% sub-sample are shown in Figure S1 and indicated that 30 iterations and 25 passes were sufficient for convergence; similar results were seen for the 0.1% sub-sample of data, and when using 50 topics. Models therefore used 30 iterations and 25 passes.

To select the optimal number of topics, we divided our data into an 90:10 train-test split and selected the number of topics resulting in the lowest value of the perplexity score on a test set.^10^ For disease sequences (Figure S2), we ran models over a range of topics from five to 100 and for Medcode sequences (Figure S3), from five to 300.^10,11^ For diseases, we selected 70 topics as optimal, and for Medcodes, selected 100 topics as optimal. Using the optimal number of topics, we then repeated the LDA algorithm using the full dataset to generate topic distributions for the full patient cohort.

**Figure S1: Trial of passes and iterations on 1% sample of data with LDA**

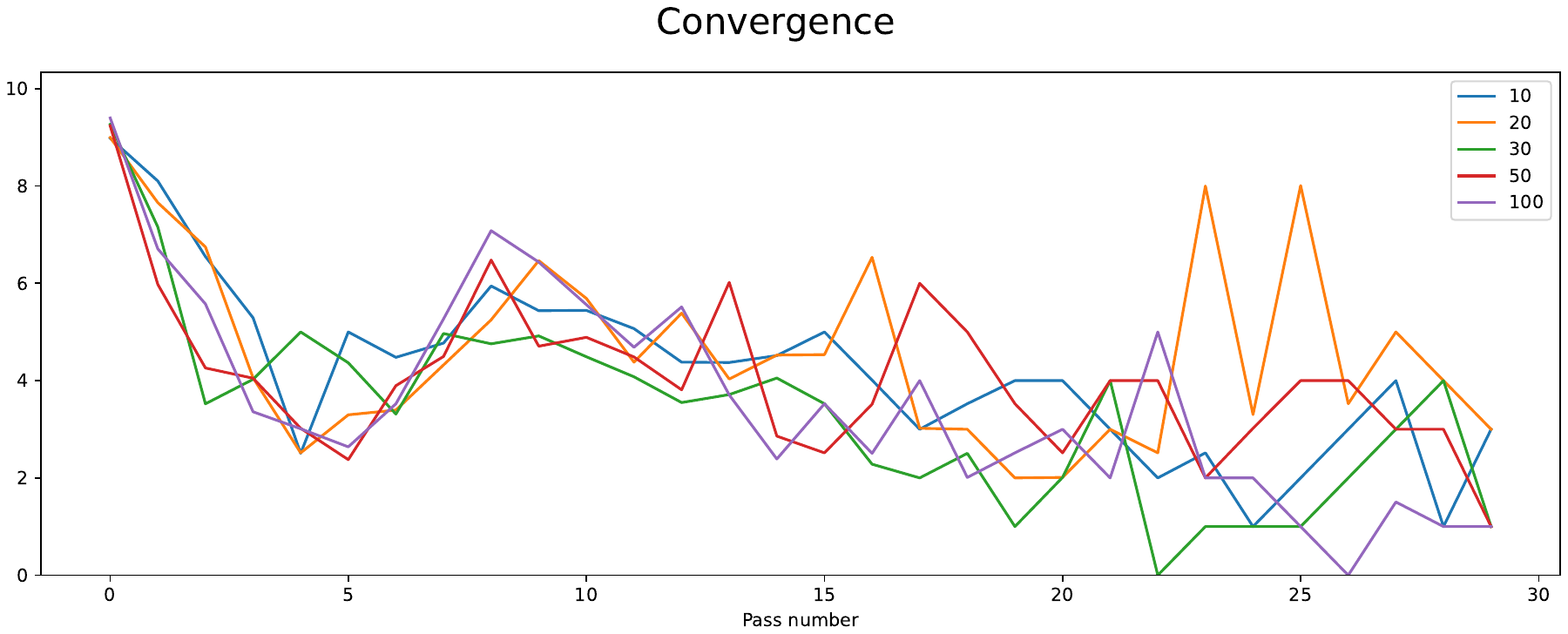

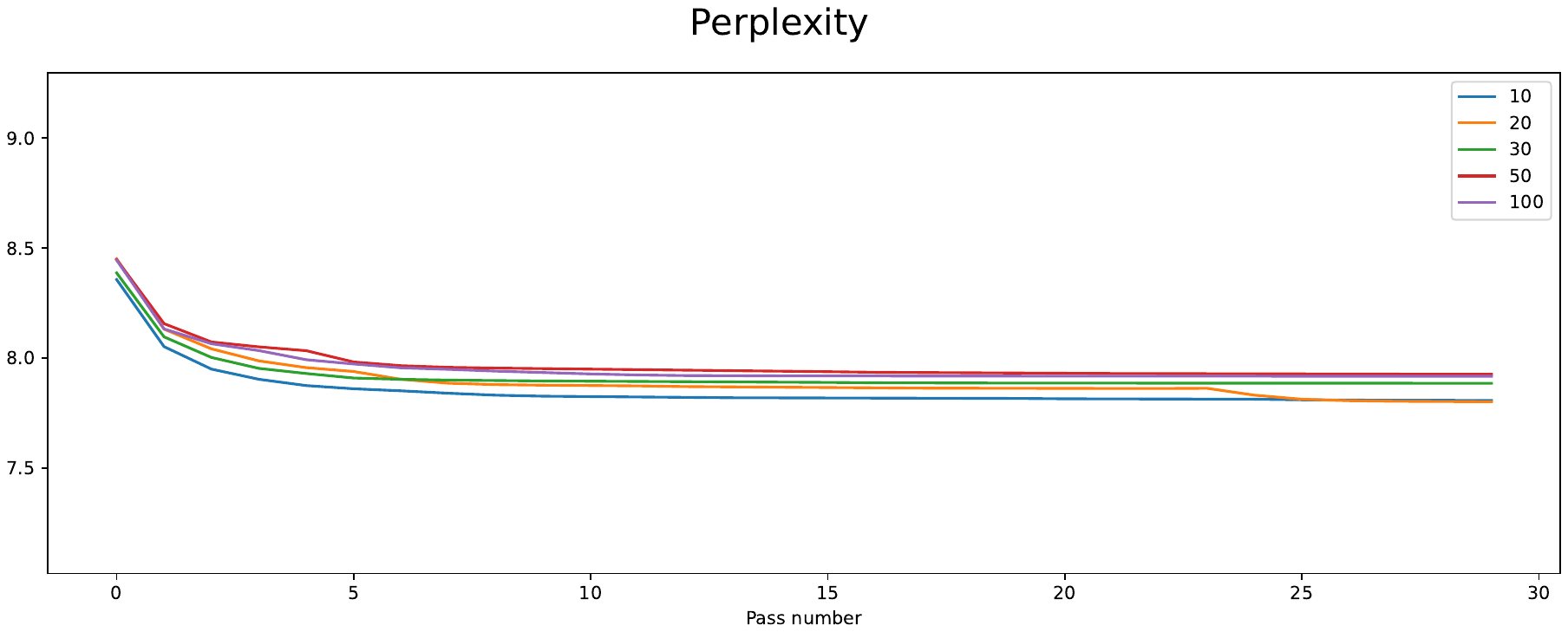

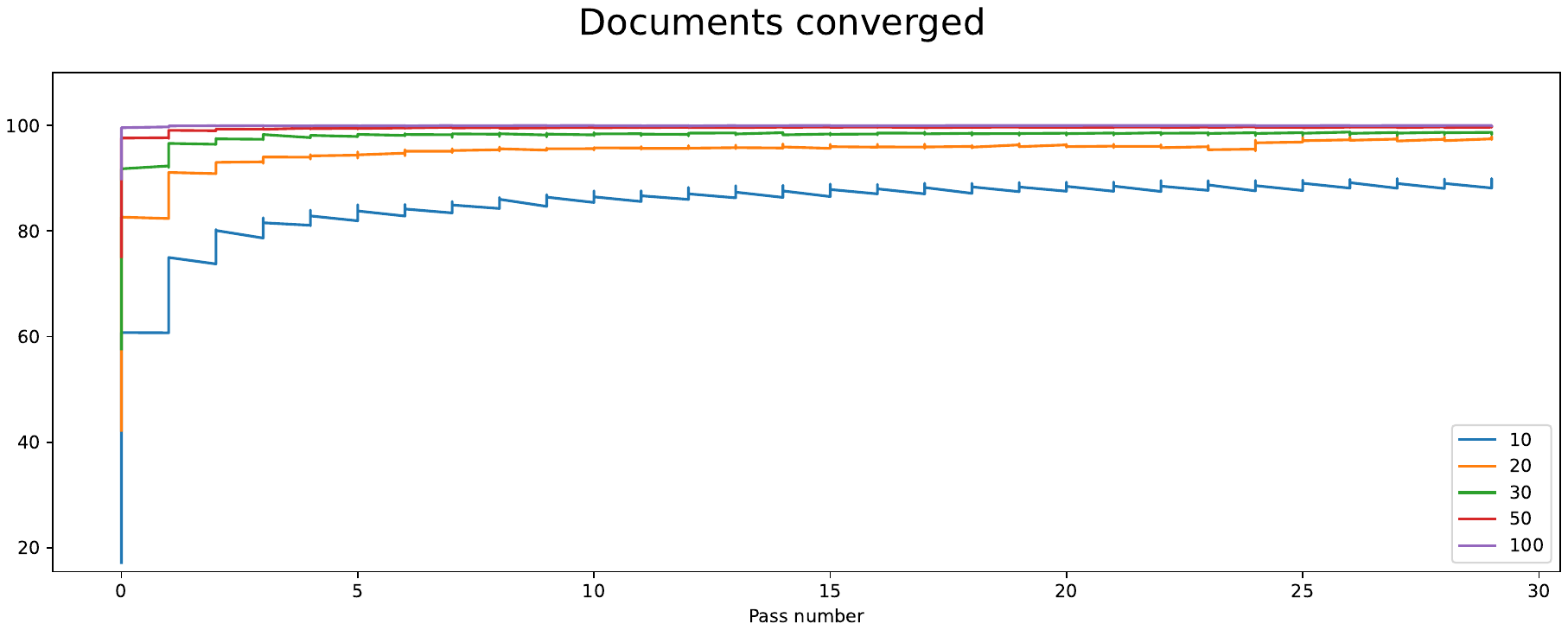

Note: y-axis displays metric values. Line colours represent number of iterations

**Figure S2: Topic perplexity on the test set for diseases**

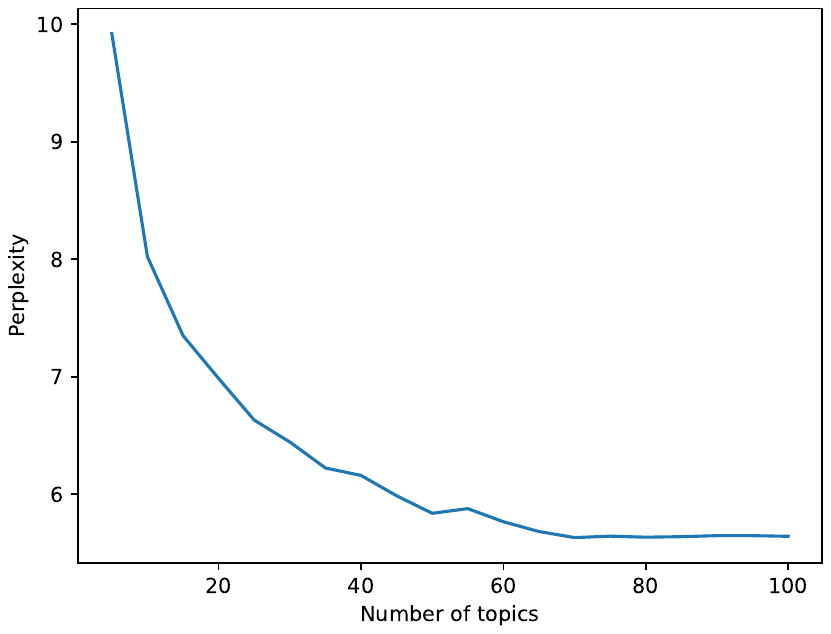

**Figure S3: Topic perplexity on the test set for Medcodes**

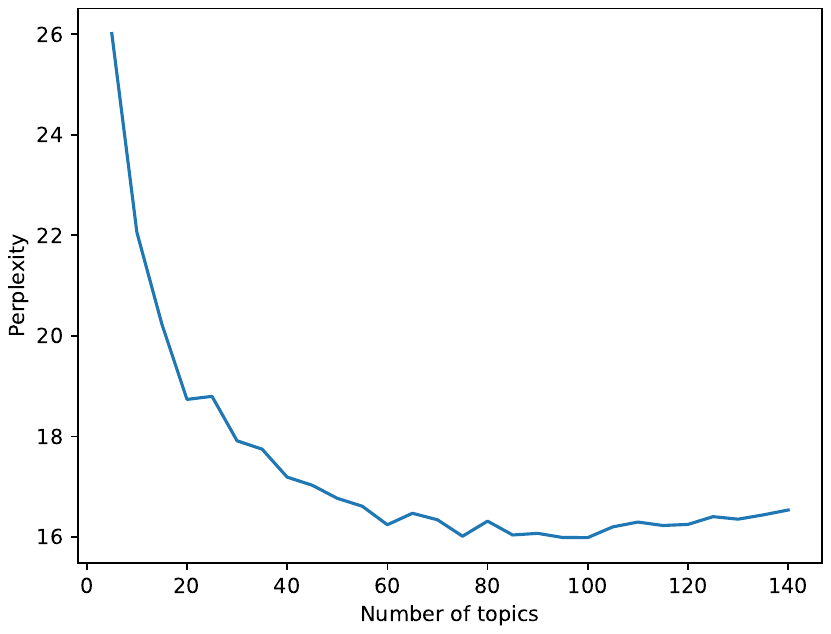

**Doc2vec**

Given our previous work using word2vec found that changing the default parameters of window size (=5), negative sampling (=5) and down-sampling (=0.001) made little difference to the learned disease representations, we retained these default settings for doc2vec.^12^ However, initial evaluations of the embeddings suggested poor results, related to the learning rate and epochs. To evaluate different hyperparameter settings of the doc2vec models, we created a metric based on patients with the same disease sequences, expecting that a well-performing model should result in patients with the same disease sequence being assigned a similar vector. We first define the set of all 1,498,469 patients with a duplicate disease sequence, *S* (with 171,378 unique disease sequences). The algorithm proceeds as follows:

1. Draw a random bootstrap sample of 10,000 patients from set *S*.
2. For each patient $P_{i}$ in set *S,* determine $D_{i}$, the sub-set of patients from set S with the same disease sequence.
3. For each patient, $P_{i}$, calculate the set ${Sim}_{i,N}$, of the *N* most similar patients based on cosine similarity from set *S*.
4. Calculate the metric $M_{i}$ as the percentage of set $D_{i}$ that are in the set ${Sim}_{i,N}$
5. Calculate the metric $M$of the mean of values of $M_{i}$ across all patients $P_{i}$

We compare values of *N* most-similar patients for 1% and 0.1% of the total dataset. The resulting metric can then be interpreted as the probability that the algorithm identifies patients which have the same disease sequence as being in the top 1% or 0.1% of most similar patients. Using this metric as a guide, experiments were run varying the learning rate, epochs and vector size, aiming to optimise the metric for values of 1% and 0.1%. For disease sequences, we trialled vector sizes of 30 and 100, learning rates of 0.025 (the default), 0.01, 0.0075, 0.005, 0.00025 and 0.001, and epochs of 10, 20, 50 and 100, shown in Table S3.

We followed the same approach for testing doc2vec models trained on Medcode sequences, shown in Table S4. Given models were well-performing using default settings, we trialled only over vector sizes of 30 and 100, learning rates of 0.025 and 0.0075 and epochs of 10, 20, 50 and 100.

**Table S3: Comparisons of doc2vec algorithms, vector size, learning rate and epochs for disease sequences**

| **Model** | **Vector size** | **Learning rate** | **Epochs** | **% exact duplicates in top 1%** | **% of exact duplicates in top 0.1%** |
| --- | --- | --- | --- | --- | --- |
| DBOW | 30 | 0.025 | 10 | 22.3% | 8.6% |
| DBOW | 30 | 0.025 | 20 | 27.8% | 12.1% |
| DBOW | 30 | 0.025 | 50 | 33.7% | 15.9% |
| DBOW | 30 | 0.025 | 100 | 35.8% | 17.8% |
| DBOW | 30 | 0.01 | 10 | 49.9% | 22.4% |
| DBOW | 30 | 0.01 | 20 | 64.7% | 35.8% |
| DBOW | 30 | 0.01 | 50 | 75.4% | 47.5% |
| DBOW | 30 | 0.01 | 100 | 78.7% | 53.5% |
| DBOW | 30 | 0.0075 | 10 | 61.7% | 31.3% |
| DBOW | 30 | 0.0075 | 20 | 81.8% | 50.3% |
| DBOW | 30 | 0.0075 | 50 | 93.5% | 69.8% |
| DBOW | 30 | 0.0075 | 100 | 96.1% | 76.1% |
| DBOW | 30 | 0.005 | 10 | 63.4% | 34.0% |
| DBOW | 30 | 0.005 | 20 | 84.3% | 56.4% |
| DBOW | 30 | 0.005 | 50 | 96.0% | 77.1% |
| DBOW | 30 | 0.005 | 100 | 98.4% | 83.6% |
| DBOW | 30 | 0.0025 | 10 | 47.4% | 20.6% |
| DBOW | 30 | 0.0025 | 20 | 77.5% | 49.0% |
| DBOW | 30 | 0.0025 | 50 | 95.1% | 75.0% |
| DBOW | 30 | 0.0025 | 100 | 98.2% | 82.9% |
| DBOW | 30 | 0.001 | 10 | 7.2% | 1.8% |
| DBOW | 30 | 0.001 | 20 | 41.1% | 16.9% |
| DBOW | 30 | 0.001 | 50 | 88.1% | 63.1% |
| DBOW | 30 | 0.001 | 100 | 96.6% | 78.4% |
| DBOW | 100 | 0.025 | 10 | 34.9% | 13.4% |
| DBOW | 100 | 0.025 | 20 | 40.7% | 17.8% |
| DBOW | 100 | 0.025 | 50 | 44.2% | 21.6% |
| DBOW | 100 | 0.025 | 100 | 46.4% | 22.9% |
| DBOW | 100 | 0.01 | 10 | 64.1% | 31.4% |
| DBOW | 100 | 0.01 | 20 | 78.5% | 44.6% |
| DBOW | 100 | 0.01 | 50 | 86.2% | 58.0% |
| DBOW | 100 | 0.01 | 100 | 88.9% | 63.2% |
| DBOW | 100 | 0.0075 | 10 | 75.2% | 40.9% |
| DBOW | 100 | 0.0075 | 20 | 89.1% | 58.5% |
| DBOW | 100 | 0.0075 | 50 | 95.3% | 74.0% |
| DBOW | 100 | 0.0075 | 100 | 96.3% | 77.7% |
| DBOW | 100 | 0.005 | 10 | 76.8% | 46.2% |
| DBOW | 100 | 0.005 | 20 | 91.7% | 65.8% |
| DBOW | 100 | 0.005 | 50 | 98.2% | 82.0% |
| DBOW | 100 | 0.005 | 100 | 99.3% | 86.2% |
| DBOW | 100 | 0.0025 | 10 | 72.4% | 41.3% |
| DBOW | 100 | 0.0025 | 20 | 90.6% | 65.3% |
| DBOW | 100 | 0.0025 | 50 | 98.2% | 82.1% |
| **DBOW** | **100** | **0.0025** | **100** | **99.4%** | **87.2%** |
| DBOW | 100 | 0.001 | 10 | 25.2% | 7.2% |
| DBOW | 100 | 0.001 | 20 | 76.9% | 45.2% |
| DBOW | 100 | 0.001 | 50 | 97.3% | 78.8% |
| DBOW | 100 | 0.001 | 100 | 99.2% | 85.8% |
| DM | 30 | 0.025 | 10 | 4.0% | 1.2% |
| DM | 30 | 0.025 | 20 | 11.6% | 5.1% |
| DM | 30 | 0.025 | 50 | 26.7% | 12.8% |
| DM | 30 | 0.025 | 100 | 45.8% | 22.0% |
| DM | 30 | 0.01 | 10 | 1.8% | 0.2% |
| DM | 30 | 0.01 | 20 | 3.1% | 0.4% |
| DM | 30 | 0.01 | 50 | 20.6% | 9.9% |
| DM | 30 | 0.01 | 100 | 53.0% | 28.1% |
| DM | 30 | 0.0075 | 10 | 1.5% | 0.1% |
| DM | 30 | 0.0075 | 20 | 2.6% | 0.5% |
| DM | 30 | 0.0075 | 50 | 15.6% | 6.5% |
| DM | 30 | 0.0075 | 100 | 50.4% | 27.4% |
| DM | 30 | 0.005 | 10 | 1.4% | 0.1% |
| DM | 30 | 0.005 | 20 | 1.9% | 0.4% |
| DM | 30 | 0.005 | 50 | 3.5% | 0.5% |
| DM | 30 | 0.005 | 100 | 8.2% | 2.7% |
| DM | 30 | 0.0025 | 10 | 1.3% | 0.1% |
| DM | 30 | 0.0025 | 20 | 1.4% | 0.1% |
| DM | 30 | 0.0025 | 50 | 1.8% | 0.3% |
| DM | 30 | 0.0025 | 100 | 2.9% | 0.5% |
| DM | 30 | 0.001 | 10 | 1.0% | 0.1% |
| DM | 30 | 0.001 | 20 | 1.0% | 0.2% |
| DM | 30 | 0.001 | 50 | 1.3% | 0.2% |
| DM | 30 | 0.001 | 100 | 1.6% | 0.3% |
| DM | 100 | 0.025 | 10 | 10.1% | 4.5% |
| DM | 100 | 0.025 | 20 | 17.8% | 8.7% |
| DM | 100 | 0.025 | 50 | 28.1% | 14.2% |
| DM | 100 | 0.025 | 100 | 33.8% | 17.9% |
| DM | 100 | 0.01 | 10 | 6.4% | 2.6% |
| DM | 100 | 0.01 | 20 | 16.6% | 8.4% |
| DM | 100 | 0.01 | 50 | 38.0% | 18.8% |
| DM | 100 | 0.01 | 100 | 54.2% | 30.0% |
| DM | 100 | 0.0075 | 10 | 4.8% | 1.6% |
| DM | 100 | 0.0075 | 20 | 12.4% | 6.1% |
| DM | 100 | 0.0075 | 50 | 31.7% | 15.6% |
| DM | 100 | 0.0075 | 100 | 44.9% | 25.1% |
| DM | 100 | 0.005 | 10 | 2.9% | 0.7% |
| DM | 100 | 0.005 | 20 | 7.4% | 3.2% |
| DM | 100 | 0.005 | 50 | 28.3% | 14.0% |
| DM | 100 | 0.005 | 100 | 32.6% | 16.6% |
| DM | 100 | 0.0025 | 10 | 1.9% | 0.2% |
| DM | 100 | 0.0025 | 20 | 3.9% | 1.2% |
| DM | 100 | 0.0025 | 50 | 13.4% | 6.8% |
| DM | 100 | 0.0025 | 100 | 34.1% | 17.7% |
| DM | 100 | 0.001 | 10 | 1.0% | 0.1% |
| DM | 100 | 0.001 | 20 | 1.3% | 0.2% |
| DM | 100 | 0.001 | 50 | 6.2% | 2.2% |
| DM | 100 | 0.001 | 100 | 14.6% | 7.0% |

* Rows highlighted in bold are those selected with highest values of both metrics

**Table S4: Comparisons of doc2vec algorithms, vector size, learning rate and epochs for Medcode sequences**

| **Model** | **Vector size** | **Learning rate** | **Epochs** | **% exact duplicates in top 1%** | **% of exact duplicates in top 0.1%** |
| --- | --- | --- | --- | --- | --- |
| DBOW | 30 | 0.025 | 10 | 60.4% | 36.8% |
| DBOW | 30 | 0.025 | 20 | 70.0% | 45.1% |
| DBOW | 30 | 0.025 | 50 | 77.5% | 54.6% |
| DBOW | 30 | 0.025 | 100 | 80.3% | 58.5% |
| DBOW | 30 | 0.0075 | 10 | 83.3% | 66.1% |
| DBOW | 30 | 0.0075 | 20 | 92.6% | 79.4% |
| DBOW | 30 | 0.0075 | 50 | 96.9% | 86.4% |
| DBOW | 30 | 0.0075 | 100 | 98.9% | 89.8% |
| DBOW | 100 | 0.025 | 10 | 80.9% | 58.9% |
| DBOW | 100 | 0.025 | 20 | 83.6% | 62.5% |
| DBOW | 100 | 0.025 | 50 | 85.6% | 67.0% |
| DBOW | 100 | 0.025 | 100 | 86.6% | 66.4% |
| DBOW | 100 | 0.0075 | 10 | 90.8% | 76.7% |
| DBOW | 100 | 0.0075 | 20 | 96.1% | 84.6% |
| DBOW | 100 | 0.0075 | 50 | 99.4% | 90.0% |
| **DBOW** | **100** | **0.0075** | **100** | **100.0%** | **92.1%** |
| DM | 30 | 0.025 | 10 | 18.2% | 7.6% |
| DM | 30 | 0.025 | 20 | 63.6% | 42.4% |
| DM | 30 | 0.025 | 50 | 92.8% | 79.4% |
| DM | 30 | 0.025 | 100 | 97.8% | 87.2% |
| DM | 30 | 0.0075 | 10 | 1.7% | 0.4% |
| DM | 30 | 0.0075 | 20 | 3.5% | 1.4% |
| DM | 30 | 0.0075 | 50 | 50.0% | 31.8% |
| DM | 30 | 0.0075 | 100 | 90.0% | 78.0% |
| DM | 100 | 0.025 | 10 | 48.5% | 34.3% |
| DM | 100 | 0.025 | 20 | 78.3% | 61.7% |
| DM | 100 | 0.025 | 50 | 93.9% | 75.9% |
| DM | 100 | 0.025 | 100 | 98.0% | 82.3% |
| DM | 100 | 0.0075 | 10 | 15.8% | 7.6% |
| DM | 100 | 0.0075 | 20 | 47.3% | 33.3% |
| DM | 100 | 0.0075 | 50 | 87.6% | 76.3% |
| DM | 100 | 0.0075 | 100 | 96.0% | 85.6% |

* Rows highlighted in bold are those selected with highest values of both metrics

**BERT models**

Figure S4 shows our model input set up for EHR-BERT. Multiple diseases/Medcodes (D1, D2, D3, etc.) can be recorded at each visit. Each visit is represented by the visit number (ordered from 1 to N), and calendar year of the visit. Additional demographic information includes age at each visit, and time-invariant attributes of gender, ethnicity, and deprivation decile. Gender was included as three categories of ‘female’, ‘male’ or ‘indeterminate/unknown’. Ethnicity was included as White, South Asian Black, Other, Mixed, or unknown. Deprivation used deciles from 1 (most deprived) to 10 (least deprived) or ‘unknown’. Note that in EHR-BERT, as with BEHRT, there is no absolute positional embedding, as there is no natural ordering of the codes within-visit, and so the absolute position is non-informative.

**Figure S4: EHR-BERT embedding model for an example of a sequence of four visits in one patient**

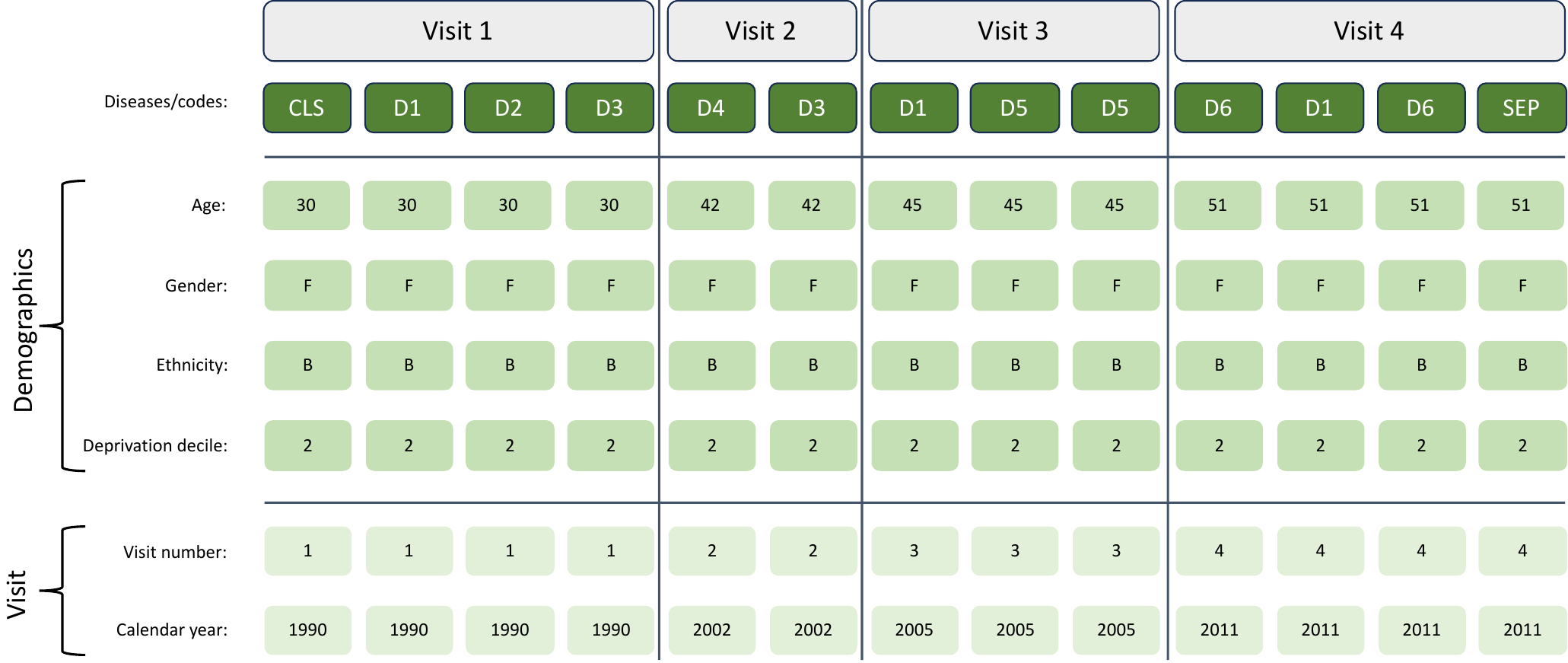

**Table S5: Comparison of prediction of mortality over different epochs for transformer models (best performance on AUC and APS for each model highlighted in bold)**

| **Model** | **Epochs** | **Covariates** | **AUC** | **APS** |
| --- | --- | --- | --- | --- |
| MedBERT diseases | 10 | Included | 0.848 | 0.182 |
| MedBERT diseases | 50 | Included | 0.848 | 0.183 |
| **MedBERT diseases** | **100** | **Included** | **0.849** | **0.184** |
| MedBERT Medcodes | 10 | Included | 0.845 | 0.179 |
| **MedBERT Medcodes*** | **50** | **Included** | **0.847** | **0.182** |
| **MedBERT Medcodes*** | **100** | **Included** | **0.847** | **0.182** |
| BEHRT diseases | 10 | Included | 0.858 | 0.205 |
| BEHRT diseases | 50 | Included | 0.860 | 0.209 |
| **BEHRT diseases** | **100** | **Included** | **0.861** | **0.210** |
| BEHRT Medcodes | 10 | Included | 0.856 | 0.202 |
| **BEHRT Medcodes*** | **50** | **Included** | **0.860** | **0.208** |
| **BEHRT Medcodes*** | **100** | **Included** | **0.860** | **0.208** |
| EHR-BERT diseases | 10 | Included | 0.864 | 0.211 |
| EHR-BERT diseases | 50 | Included | 0.866 | 0.215 |
| **EHR-BERT diseases** | **100** | **Included** | **0.866** | **0.216** |
| EHR-BERT Medcodes | 10 | Included | 0.860 | 0.206 |
| EHR-BERT Medcodes | 50 | Included | 0.863 | 0.210 |
| **EHR-BERT Medcodes** | **100** | **Included** | **0.863** | **0.211** |

***** Indicates where best model not consistent between AUC and APS (one model results in the highest AUC, another in the highest APS)

**Table S6: Comparison of prediction of any hypertension code over different epochs for transformer models (best performance on AUC and APS for each model highlighted in bold)**

| **Model** | **Epochs** | **Covariates** | **AUC** | **APS** |
| --- | --- | --- | --- | --- |
| MedBERT diseases | 10 | Included | 0.915 | 0.559 |
| MedBERT diseases | 50 | Included | 0.915 | 0.558 |
| **MedBERT diseases** | **100** | **Included** | **0.915** | **0.562** |
| MedBERT Medcodes | 10 | Included | 0.912 | 0.554 |
| **MedBERT Medcodes*** | **50** | **Included** | **0.912** | **0.555** |
| **MedBERT Medcodes*** | **100** | **Included** | **0.912** | **0.556** |
| BEHRT diseases | 10 | Included | 0.920 | 0.592 |
| BEHRT diseases | 50 | Included | 0.920 | 0.600 |
| **BEHRT diseases** | **100** | **Included** | **0.920** | **0.606** |
| BEHRT Medcodes | 10 | Included | 0.917 | 0.581 |
| BEHRT Medcodes | 50 | Included | 0.918 | 0.593 |
| **BEHRT Medcodes** | **100** | **Included** | **0.918** | **0.598** |
| EHR-BERT diseases | 10 | Included | 0.921 | 0.605 |
| EHR-BERT diseases | 50 | Included | 0.920 | 0.613 |
| **EHR-BERT diseases** | **100** | **Included** | **0.920** | **0.616** |
| EHR-BERT Medcodes | 10 | Included | 0.915 | 0.580 |
| EHR-BERT Medcodes | 50 | Included | 0.917 | 0.596 |
| **EHR-BERT Medcodes** | **100** | **Included** | **0.918** | **0.602** |

***** Indicates where best model not consistent between AUC and APS (one model results in the highest AUC, another in the highest APS)

**Supplementary results**

**Figure S5: Model ROC-AUC (panel A) and APS (panel B) for different embedding models for prediction of mortality, emergency department attendances and emergency admissions within 12 months; without covariates)**

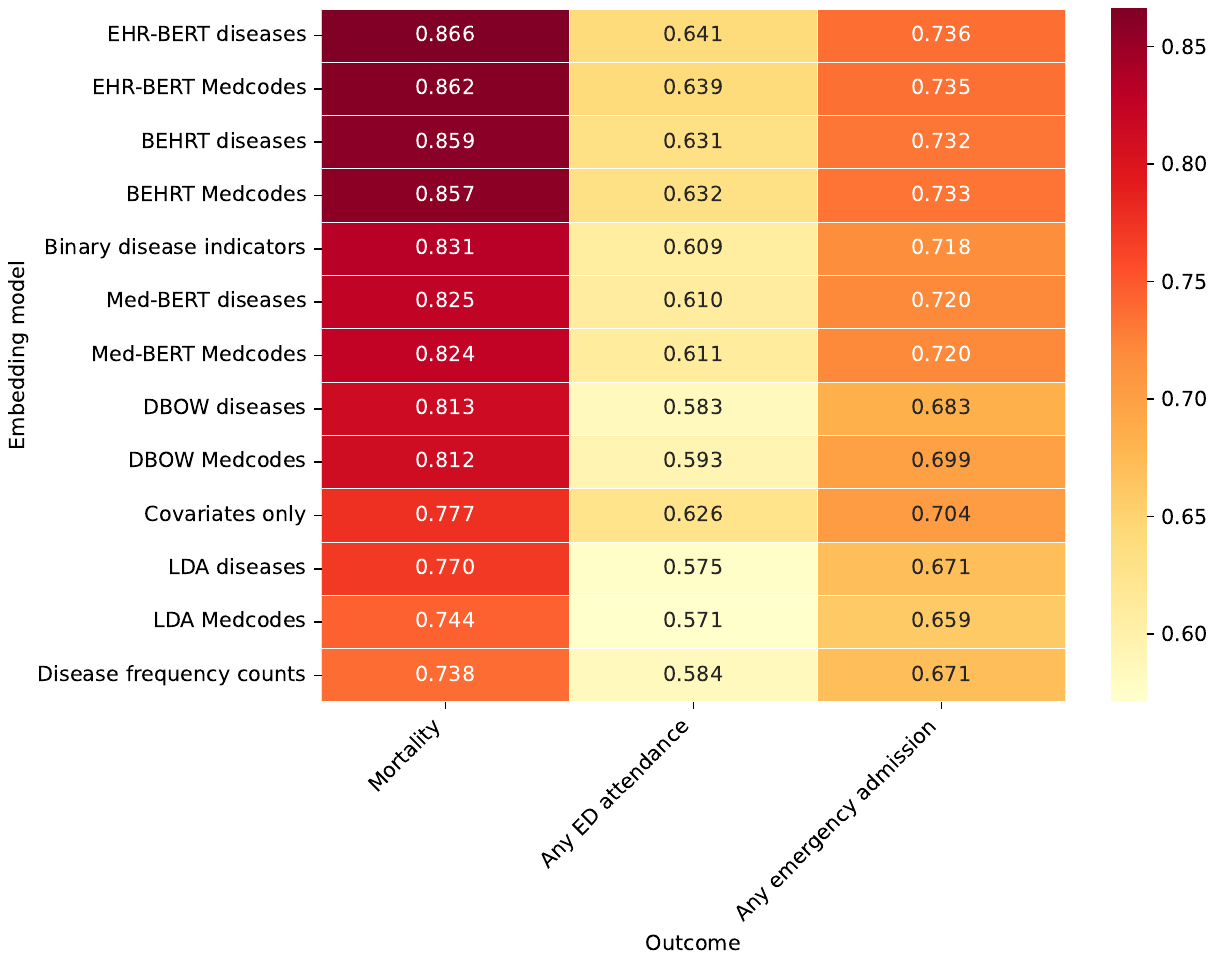

**A**

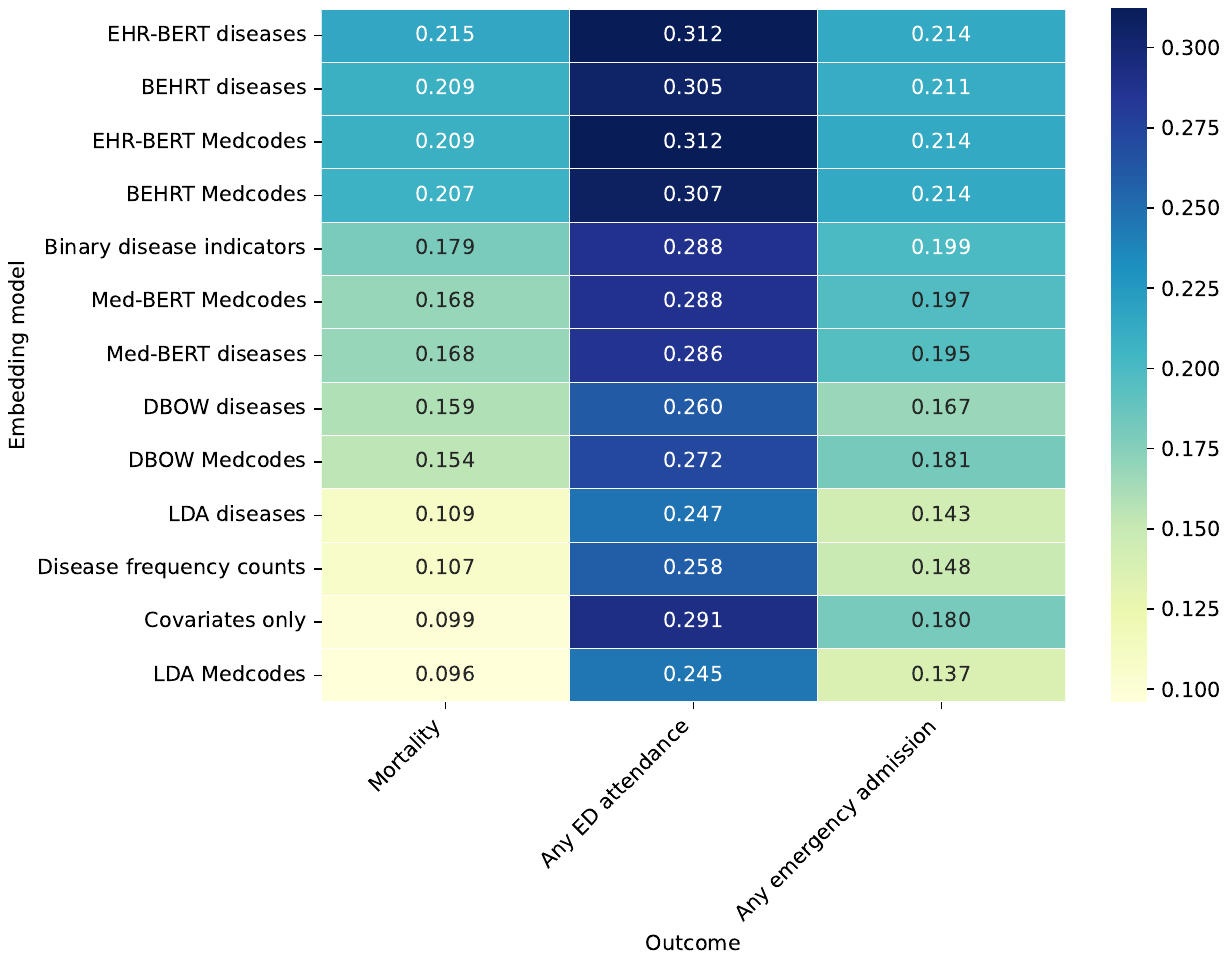

**B**

**Figure S6: Model ROC-AUC (panel A) and APS (panel B) for different embedding models for prediction of any versus new diseases developed within 12 months; without covariates**

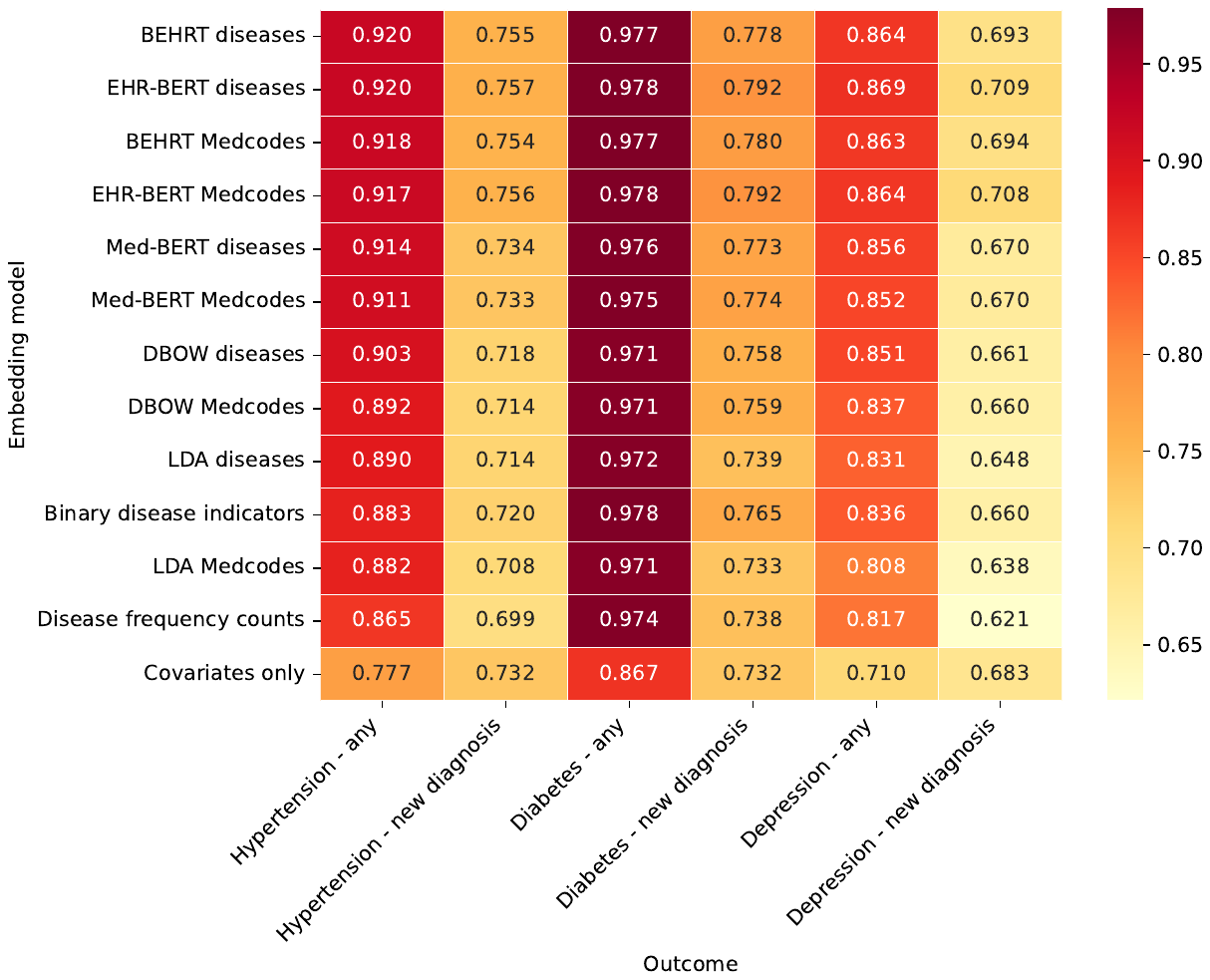

**A**

**B**

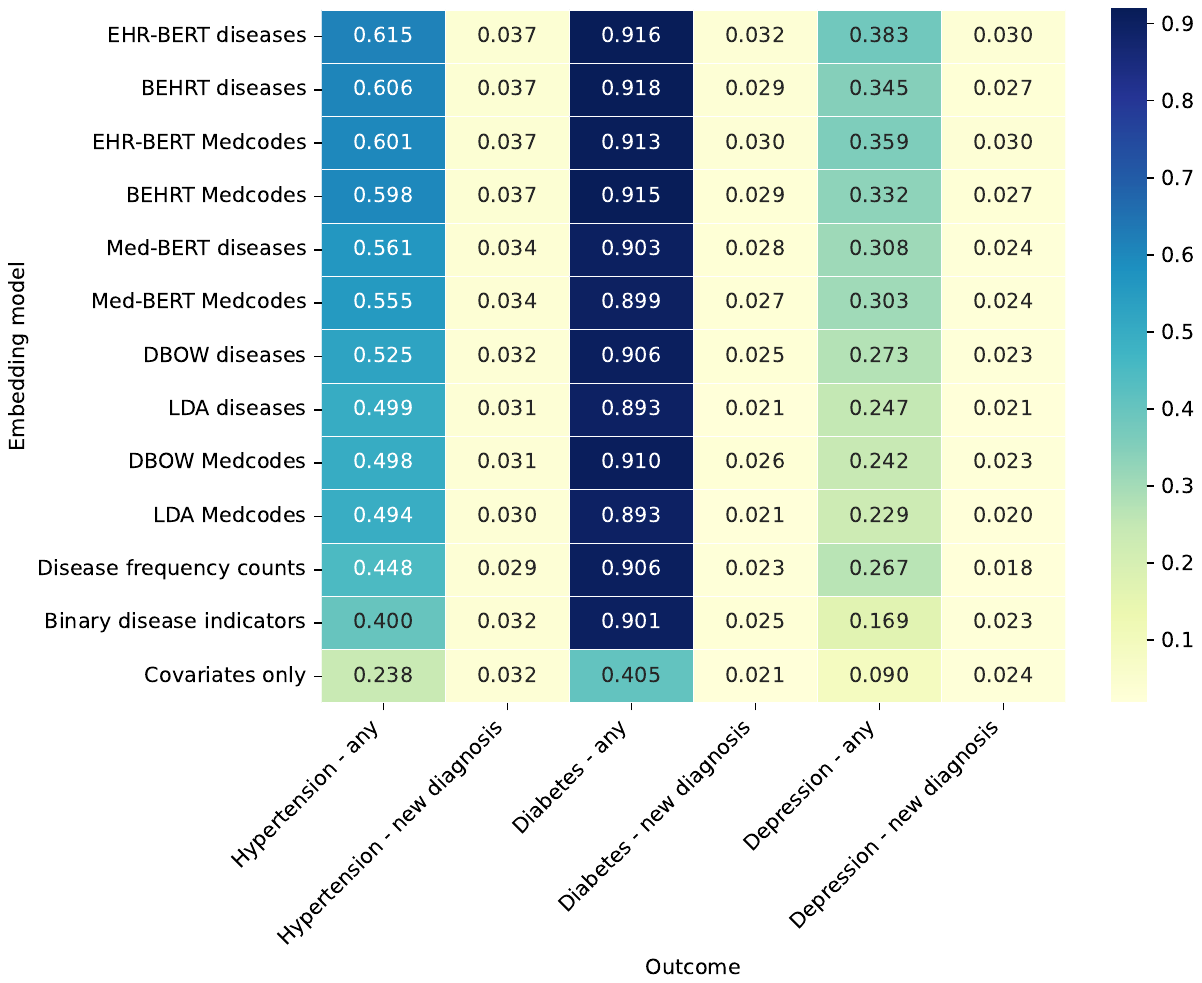

**References**

1.	NHS Digital. Data Model and Dictionary: Ethnic Category. https://www.datadictionary.nhs.uk/data_elements/ethnic_category.html.

2.	Davidson, J. *et al.* Codelists for: ‘Ethnic differences in the incidence of clinically diagnosed influenza: an England population-based cohort study 2008-2018’. https://datacompass.lshtm.ac.uk/id/eprint/2102/ (2021) doi:10.17037/DATA.00002102.

3.	Mathur, R., Palla, L., Farmer, R. E., Chaturvedi, N. & Smeeth, L. Ethnic differences in the severity and clinical management of type 2 diabetes at time of diagnosis: A cohort study in the UK Clinical Practice Research Datalink. *Diabetes Research and Clinical Practice* **160**, (2020).

4.	Gallagher, A. M., Dedman, D., Padmanabhan, S., Leufkens, H. G. M. & de Vries, F. The accuracy of date of death recording in the Clinical Practice Research Datalink GOLD database in England compared with the Office for National Statistics death registrations. *Pharmacoepidemiology and Drug Safety* **28**, 563–569 (2019).

5.	Delmestri, A. & Prieto-Alhambra, D. CPRD GOLD and linked ONS mortality records: Reconciling guidelines. *Int J Med Inform* **136**, 104038 (2020).

6.	Wallach, H., Mimno, D. & McCallum, A. Rethinking LDA: Why Priors Matter. in *Advances in Neural Information Processing Systems* vol. 22 (Curran Associates, Inc., 2009).

7.	Syed, S. & Spruit, M. Selecting Priors for Latent Dirichlet Allocation. in *2018 IEEE 12th International Conference on Semantic Computing (ICSC)* 194–202 (IEEE, 2018). doi:10.1109/ICSC.2018.00035.

8.	Hoffman, M., Bach, F. & Blei, D. Online Learning for Latent Dirichlet Allocation. in *Advances in Neural Information Processing Systems* (eds. Lafferty, J., Williams, C., Shawe-Taylor, J., Zemel, R. & Culotta, A.) vol. 23 (Curran Associates, Inc., 2010).

9.	Řehůřek, R. & Sojka, P. Software Framework for Topic Modelling with Large Corpora. in *Proceedings of the LREC 2010 Workshop on New Challenges for NLP Frameworks* 45–50 (ELRA, 2010).

10.	Röder, M., Both, A. & Hinneburg, A. Exploring the Space of Topic Coherence Measures. in *Proceedings of the Eighth ACM International Conference on Web Search and Data Mining* 399–408 (Association for Computing Machinery, 2015). doi:10.1145/2684822.2685324.

11.	Stevens, K., Kegelmeyer, P., Andrzejewski, D. & Buttler, D. Exploring Topic Coherence over Many Models and Many Topics. in *Proceedings of the 2012 Joint Conference on Empirical Methods in Natural Language Processing and Computational Natural Language Learning* 952–961 (Association for Computational Linguistics, 2012).

12.	Beaney, T. *et al.* Identifying multi-resolution clusters of diseases in ten million patients with multimorbidity in primary care in England. 2023.06.30.23292080 Preprint at https://doi.org/10.1101/2023.06.30.23292080 (2023).
